# Prediction of depressive symptoms severity based on sleep quality, anxiety, and brain: a machine learning approach across three cohorts

**DOI:** 10.1101/2023.08.09.23293887

**Authors:** Mahnaz Olfati, Fateme Samea, Shahrooz Faghihroohi, Somayeh Maleki Balajoo, Vincent Küppers, Sarah Genon, Kaustubh Patil, Simon B. Eickhoff, Masoud Tahmasian

## Abstract

**Background:** Depressive symptoms are rising in the general population, but their associated factors are unclear. Although the link between sleep disturbances and depressive symptoms severity (DSS) is reported, the predictive role of sleep on DSS and the impact of anxiety and the brain on their relationship remained obscure.

**Method:** Using three population-based datasets, we trained the machine learning models in the primary dataset (N = 1101) to assess the predictive role of sleep quality, anxiety, and brain structure and function measurements on DSS, then we tested our models’ performance in two independent datasets (N = 334, N = 378) to test the generalizability of our findings. Furthermore, we applied our machine learning model to a smaller longitudinal sample (N = 66). In addition, we performed a mediation analysis to identify the role of anxiety and brain measurements on the sleep quality-DSS link.

**Findings:** Sleep quality could predict individual DSS (r = 0.43, R^2^ = 0.18, rMSE = 2.73), and adding anxiety, rather than brain measurements, strengthened its prediction performance (r = 0.67, R^2^ = 0.45, rMSE = 2.25). Importantly, out-of-cohort validations in other cross-sectional datasets and a longitudinal sample provided robust results. Furthermore, anxiety scores (not brain measurements) mediated the association between sleep quality and DSS.

**Interpretation:** Poor sleep quality could predict DSS at the individual subject level across three cohorts. Anxiety symptoms not only increased the performance of the predictive model but also mediated the link between sleep and DSS.

**Research in Context:** *Evidence before this study:* Depressive symptoms are prevalent in modern societies, but their associated factors are less identified. Several studies suggested that sleep disturbance and anxiety are linked with depressive problems in the general population and patients with major depressive disorder. A few longitudinal studies and meta-analyses also suggested that sleep disturbance plays a key role in developing depressive problems and clinical depression. However, those original studies mainly used conventional group comparison statistical approaches, ignoring the inter-individual variability across participants. Moreover, their data were limited to a single cohort, limiting the generalizability of their findings in other samples. Thus, large-scale multi-cohort studies using machine learning predictive approaches are needed to identify the complex relationship between sleep quality, anxiety symptoms, and depressive symptoms at the individual subject level. We also focused on the neurobiological underpinning of their interplay.

*Added value of this study:* In this study, we used machine learning which enables individual-level predictions and can validate models on unseen data, thus providing a more robust analytical framework. This study used three independent cohorts, included a longitudinal sample, and performed careful complementary analyses to examine the robustness of our findings considering the impact of lifetime history of depression, effects of sleep-related questions of the depressive assessment, most important parameters of sleep quality in prediction of depressive symptoms severity, and testing the reverse direction i.e., predicting sleep quality based on depressive symptoms. We found that poor sleep quality could robustly predict depressive symptoms across three cohorts, but the reverse direction (prediction of sleep quality based on depressive symptoms) was less robust. Anxiety symptoms improved the performance of the predictive model and mediated the link between sleep and depressive symptoms. However, brain structure and function did not play an important role in their association. Our longitudinal data also highlighted the predictability of future depressive symptoms severity and the role of interventions (i.e., neurofeedback) in the prediction of future depressive symptoms based on sleep and anxiety.

*Implications of all the available evidence:* As depressive symptoms have a strong impact on public health, identifying their contributing factors such as poor sleep and anxiety is critical to decrease the burden of depressive symptoms and/or design better therapeutical approaches at the individual subject level.

## Introduction

In modern societies, about 25% of the general population presents depressive symptoms such as sadness, irritability, anhedonia, low motivation, distracted concentration, worthlessness, abnormal appetite, and sleep disturbance.^1,2^ Depressive symptoms have dramatically increased in general populations from 1991 to 2018, mainly in young women.^3^ Recent findings during the COVID-19 pandemic observed that the prevalence of depressive symptoms increased about 3-fold compared to the earlier population-based estimates of mental health.^4^ Critically, depressive symptoms could predict major depressive disorder (MDD) around 15 years later in white adults.^5^ Hence, screening subjects with depressive symptoms in the general population is essential for decreasing the rate, burden, and severity of depression.^6^ In addition, a high conversion rate of depressive symptoms to MDD ^5^ and the noticeable health-related and economic burden of depressive problems in the general population^7^ makes it imperative to identify the associated behavioral and brain factors of depressive phenotype.

Our life experiences highlight a significant mood impairment after night(s) of sleep disturbances, suggesting a robust link between poor sleep and depressive symptoms ^8–10^. In particular, several meta-analyses suggested that sleep disturbance, and particularly insomnia, are critical factors for developing depression^11–14^. Treatment of sleep problems reduces depressive symptoms and MDD,^15–17^ suggesting that targeting sleep quality is necessary for the management of depressive problems. On the other hand, insomnia/hypersomnia are among the diagnostic criteria of MDD, suggesting a bidirectional association between sleep and depression. Nevertheless, many individuals with sleep problems never develop depressive symptoms and some patients with depressive phenotype report normal sleep patterns, which makes the interrelationships between sleep disturbance and depressive profile very complex. The potential reasons could be due to inter-individual variability in emotional distress, anxiety, hyperarousal state, emotion regulation abilities, and coping strategies for stressful life events.^18–22^ The open question is whether depressive symptoms can be predicted based on sleep quality at the individual subject level and what underlying behavioral and brain factors contribute to their associations.

Anxiety is the most prominent mental condition that co-occurs with both sleep disturbance and depression.^10,23^ Moreover, a growing body of neuroimaging evidence highlighted the structural and functional brain alterations, mainly in the default mode and salience networks, on the interplay between sleep and depressive symptoms.^24^ Using the Human Connectome Project in young adults (HCP-Young) cohort, Cheng and colleagues ^25^ demonstrated that increased functional connectivity between several brain regions mediates the association between depressive symptom severity (DSS) and sleep quality. The volume of the hippocampus mediates the association between sleep quality and depressive symptoms. ^26^

Existing behavioral and neuroimaging studies on the link between sleep and depressive symptoms have used conventional statistical methods (mainly group comparisons and/or correlations) using a single cohort,^24^ which is prone to deliver poor generalizability in other samples. Thus, the “real world” challenge is a prediction of depressive symptoms in unseen data or independent samples to achieve generalization to future cases that cannot be answered in traditional statistical approaches based on a single sample. Advanced machine learning (ML) predictive models provide hope to identify the role of neurobehavioral factors in predicting depressive problems across various general population samples, which is crucial for precision psychiatry and ultimately guiding clinical practice.^27,28^

Thus, aiming to address these gaps in the literature, we applied the ML approach in the HCP-Young dataset to predict DSS based on sleep quality, anxiety, and the brain’s gray matter volume (GMV). In addition, we assessed the role of functional brain measurements i.e., regional homogeneity (ReHo) or fractional amplitude of low-frequency fluctuations (fALFF) in the complementary analyses. Based on the trained ML models in the HCP-Young, out-of-cohort validation of our ML algorithm was conducted on two independent US population-based datasets (i.e., the lifespan Human Connectome Project (HCP-Aging) and enhanced Nathan Kline Institute-Rockland sample (eNKI)) to understand the generalizability of our models across different cohorts. Further, we applied our models on a small set of longitudinal samples from eNKI dataset to predict future DSS. In addition, we aimed to understand the mediatory role of anxiety and GMV in the association between sleep quality and DSS in the HCP-Young dataset.

## Methods

### Cohorts

HCP-Young is a general population dataset acquired by the Washington University-University of Minnesota (WU-Minn HCP) consortium (https://www.humanconnectome.org/).^29^ Their inclusion criteria included healthy young adult (22–35 years) participants with no significant history of psychiatric disorder, substance abuse, neurological, or cardiovascular disease nor pharmaceutical or behavioral treatment. From this dataset, as our primary dataset, we included all participants who had 3T structural MRI images and phenotypic data that we were interested in in this study, i.e., sleep quality, anxiety, and depressive symptoms. In a complementary analysis, we removed participants with a lifetime history of diagnosed depression.

The HCP-Aging (https://www.humanconnectome.org/) ^30^ cohort recruited more than 1200 healthy adults aged 60 to above 100.^31^ However, we included participants aged 36 to 59 who filled out questionnaire forms since the DSS questionnaires had been designed for young and middle-aged adults. The eNKI is also a large-scale community representative dataset of the general population with cross-sectional and longitudinal samples (http://fcon_1000.projects.nitrc.org/indi/enhanced/).^32^ From this dataset, we included participants (age range 18–59 years) with cross-sectional records for predictive ML assessment and those who had longitudinal records to forecast future depression.

### Behavioral measures

#### Sleep quality

Sleep quality assessment was based on the self-reported PSQI questionnaire [27], which has 19 questions assessing sleep quality over one month. The PSQI comprises seven components, namely subjective sleep quality, sleep latency, sleep duration, habitual sleep efficiency, sleep disturbances, use of sleep medicine, and daytime dysfunction. The total PSQI score is a sum of these components. Of note, the higher total PSQI score (> 5) reflects poor sleep quality.

#### Depressive symptom severity

Depressive symptoms were measured based on the DSM-IV-oriented depressive problems portion of the Achenbach Adult Self-Report (ASR) for ages 18-59.^33^ This questionnaire has 123 items in general, and a total depressive score obtained from 14 depressive-related items, ranges from 0 to 28 points. The higher score reveals severe depressive symptoms and the sex-/age-adjusted t-score above 69 reveals the clinical range. Notably, there are two sleep-related items in this questionnaire, which have been removed in our primary ML and mediation analyses. These questions were “I sleep more than most other people during the day and/or night” and “I have trouble sleeping”. We calculated the total score of depressive problems after removing sleep-related items and used this total score in our analyses. Further, as a complementary analysis, we examined the original DSS (we refer to it as DSS’), which involves these two sleep-related items.

#### Anxiety

Anxiety score was measured using six relevant items of DSM-IV-oriented ASR for the age range 18-59. None of these items are related to sleep or depressive problems. Similar to DSS, the total score of anxiety has been used in our study and a higher anxiety score shows more anxiety problems.

### Neuroimaging measures

In this study, we used parcel-wise whole-brain GMV to assess the role of brain structure in the link between sleep quality and DSS across three cohorts. Further, we assessed resting-state fMRI features of the brain (i.e., ReHo and fALFF of the same parcels) in the HCP-Young dataset as confirmatory analyses to assess the role of the brain at the functional level (see more details in the supplementary material).

#### Calculation of gray matter parcel volume

T1 structural MRI images were acquired by Siemens 3T Skyra scanner and preprocessed using the WU-Minn HCP consortium pipelines.^34^ We performed voxel-based morphometry (VBM) using the Computational Anatomy Toolbox (CAT12),^35^ implemented in the Statistical Parametric Mapping (SPM12, https://www.fil.ion.ucl.ac.uk/spm/software/spm12/). During this process, we corrected bias-field distortions and after noise removal and skull striping, the images were normalized to standard space MNI-152. Then, we segmented the brain tissue into gray matter, white matter, and cerebrospinal fluid. Subsequently, we modulated the gray matter segments for the non-linear transformations performed during normalization to obtain the actual volumes. GMVs of the cortical, subcortical, and cerebellar areas were assessed using functionally-informed in-vivo atlases (400 cortical parcels from Schaefer atlas,^36^ 36 subcortical parcels from Brainnetome,^37^ and 37 cerebellar parcels from Buckner ^38^), resulting in 473 brain parcels, as applied previously.^39^

### Statistical analyses

#### Prediction analysis in the HCP-Young dataset

Ensemble decision tree methods were employed to structure predictive models using MATLAB R2020a software.^40^ Ensemble methods of these models were LS-boost and bagging, which were applied as a hyperparameter to be selected automatically by the algorithm (see below). First, we performed nested 10-fold cross-validation considering the family structure of subjects, in which twins and siblings were not separated in the training, validation, and test sets to avoid potential leakages. We used training sets to construct models, validation sets to select hyperparameters and feature numbers, and unseen test sets to finally evaluate the models’ performance (**Figure 1**). Then, regression models were made to regress out age, sex, and total GMV from features of training sets and subsequently, these models were used for regressing out control variables of test sets. Then, features of training sets were ranked and sorted (from the maximum importance to the minimum importance) by the relief method to enable the algorithm to select features based on the maximum rank.^41^ After putting aside the validation sets, models were constructed and trained in each remained training set ten times by ten different feature numbers so that the number of features also was able to be selected automatically based on the minimum error of prediction of the validation sets. In this stage, hyperparameters were optimized using the Bayesian method,^42^ with 100 iterations. Then, models with the minimum error of prediction of validation sets were selected and fitted on the entire training sets (training + validation) and finally used to predict unseen test sets. Thus, in the end, we had ten models (one model for each test set), and our ML pipeline could select different algorithms LS-Boost/bagging along with its hyperparameters and different feature numbers for each fold. These predictive models had 19 input features consisting of PSQI questions. Subsequently, we added anxiety (total score) and whole-brain GMV (n = 473) features to measure the role of anxiety and GMV in DSS prediction. Of note, against models with a combination of features of GMV, we did not perform a feature section for models with just sleep quality and/or anxiety features because the number of features was not too high that needed feature selection. More details of these ML analyses, hyperparameters, and feature numbers are provided in the supplementary materials.

**Figure 1.**
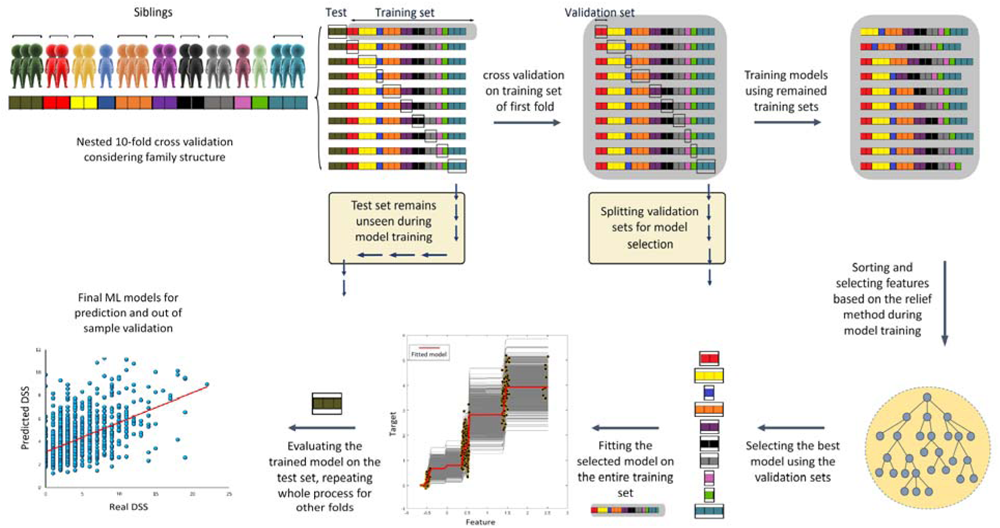
ML pipeline for prediction of DSS (depressive symptoms severity) considering family structure. First of all, 10-fold cross-validation was performed so that siblings were not separated in training/test sets. After putting aside the test set (of the first fold from now), we performed a 10-fold cross-validation on the training set (of the first fold) considering family structure. In this stage, we split validation sets and trained models on the remaining training sets. On each fold, we trained models and optimized hyper-parameters ten times with ten different feature numbers. Hence, we had ten folds and ten models for each fold and the algorithm had to select the model with the best performance and minimum error across all folds. Subsequently, the selected model was fitted on the entire training set and then evaluated on the test set. This process repeated for all other nine folds (Note: all units in the figure are arbitrary, DSS: depressive symptoms severity after excluding two sleep-related items).

### Complementary analyses in the HCP-Young dataset

In several follow-up analyses, we controlled for potential issues to examine the robustness of our findings and to cover different aspects of the interplay between behavioural and brain variables as follows: 1) we assessed correlation between sleep quality features to test feature redundancy (**eFigure 1**); 2) to test the role of functional brain scores in our main predictive ML analyses, we calculated ReHO and fALFF of 473 parcels from resting-state fMRI images in the primary database (HCP-Young) (**eFigure 2**); 3) we assessed the predictive role of anxiety (alone) and combination of GMV and anxiety features separately (**eFigure 3**); 4) in order to test multicollinearity between variables, we performed cross-prediction of anxiety and DSS and also tested collinearity between all phenotypic parameters using variance inflation factor (**eFigure 4**); 5) we removed 103 participants with a lifetime history of diagnosed depression to assess the confounding role of history of clinical depression in some individuals (**eFigure 5**); 6) we used seven components of PSQI, instead of individual PSQI items (**eFigure 6**); 7) and their feature importance (**eFigure 7**); 8) critically, in order to assess the reverse direction of prediction, we assessed the predictability of sleep quality based on depressive symptoms (**eFigure 8**), 9) we examined the predictability of sleep quality based on GMV (alone) (**eFigure 9**); 10) we also used original DSS questionnaire including two sleep-related items (of note, we deleted those items in our main analyses) (**eFigure 10**); 11) compared the results with and without sleep-related items of in the DSS questionnaire (**eFigure 11**); and 12) we tested the role of anxiety and GMV in the link between sleep quality and DSS by several mediation analyses (**eFigure 12**). Details of these complementary analyses are described in the supplementary material.

**Figure 2.**
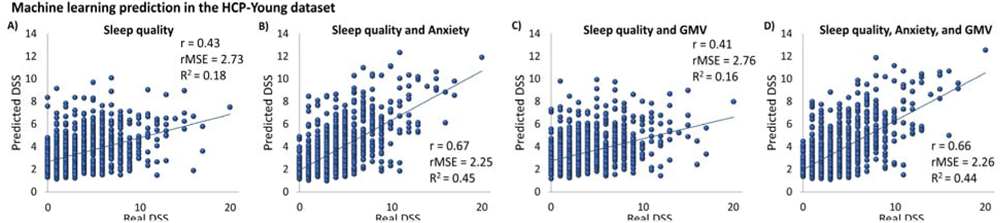
Prediction of DSS in HCP-Young dataset. A) prediction based on sleep quality; B) prediction based on a combination of sleep quality and anxiety; C) prediction based on a combination of sleep quality and GMV D) prediction based on a combination of sleep quality and anxiety and GMV (GMV: gray matter volume, DSS: depressive symptoms severity after excluding two sleep-related items, r: correlation coefficient between real and predicted DSS, rMSE: root mean squared error, R^2^: determination coefficient)

**Figure 3.**
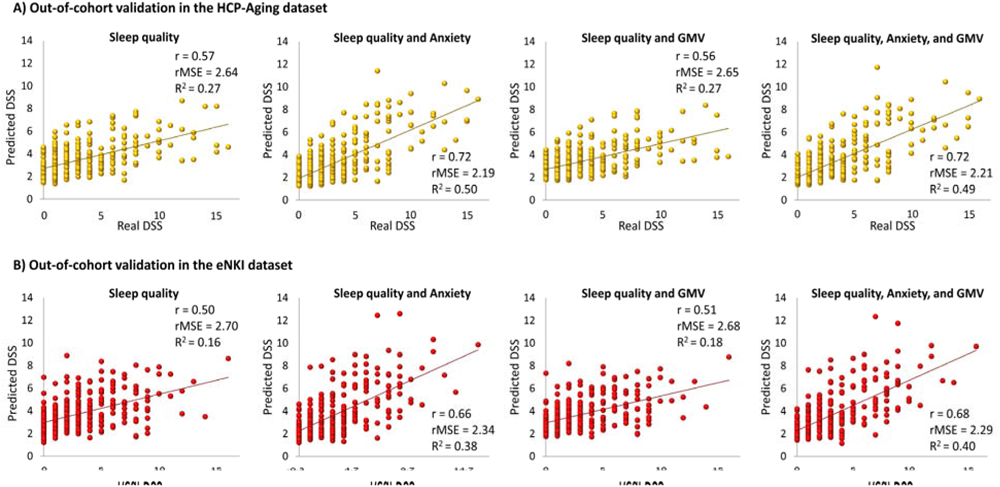
Out-of-cohort validation of ML results in two independent datasets. A) prediction of DSS in HCP-Aging dataset based on sleep quality, a combination of sleep quality and anxiety, a combination of sleep quality and GMV, a combination of sleep quality and anxiety, and GMV; B) prediction of DSS in eNKI dataset based on sleep quality, a combination of sleep quality and anxiety, a combination of sleep quality and GMV, a combination of sleep quality and anxiety, and GMV (GMV: gray matter volume, DSS: depressive symptoms severity after excluding two sleep-related items, r: correlation coefficient between real and predicted DSS, rMSE: root mean squared error, R^2^: determination coefficient)

**Figure 4.**
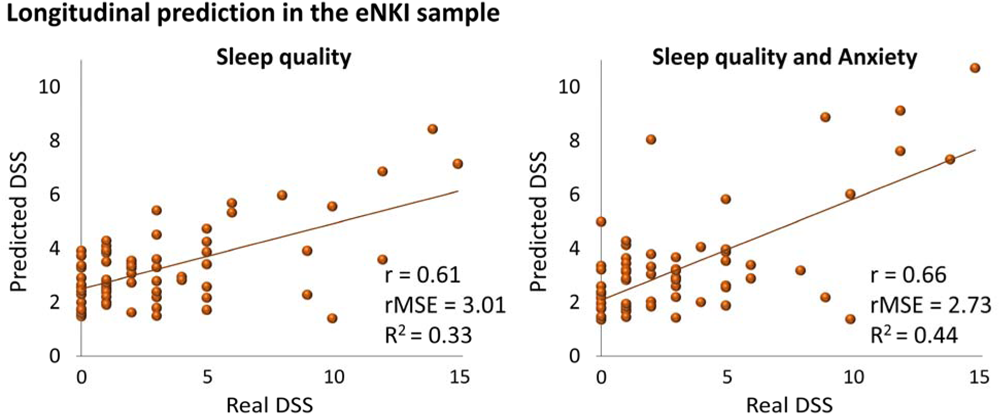
Prediction of future DSS based on sleep quality and anxiety. There were 66 participants in the longitudinal sample of the eNKI dataset (DSS: depressive symptoms severity after excluding two sleep-related items, r: correlation coefficient between real and predicted DSS, rMSE: root mean squared error, R^2^: determination coefficient)

### Out-of-cohort validation in two independent datasets

We used two independent large-scale datasets to test whether the results of ML models using the HCP-Young dataset are generalizable to other independent datasets (i.e., the eNKI and HCP-Aging). After training ML models on the HCP-Young dataset, we achieved ten models for each prediction, froze them, and used them to predict the individual DSS in other datasets and averaged the results of all ten models for each person. Of note, we did not tune our models nor performed cross-validation for these independent datasets, to keep the original model parameters steady. Put differently, we used these independent datasets solely for prediction and used the regression model of the primary dataset (HCP-Y) for regressing out age, sex, and total GMV in these datasets as well. All the phenotypic data (sleep quality, anxiety, and DSS) were obtained from the same questionnaires across the three datasets. Further, we used a small longitudinal sample of the eNKI dataset to predict future individual DSS based on present sleep quality and anxiety. We used the sleep quality and anxiety of their first records as features and the DSS of their second records as the targets. Then, we calculated the correlation between the predicted DSS and the DSS of the second record. Finally, as the complementary analyses, we separated participants who had either received or not received neurofeedback therapy intervention between their first and second visits and assessed their predictive performance to identify the role of therapeutical interventions in longitudinal predictions (**eFigure 13**).

### Mediation analysis

The structural equation modeling (SEM) using Amos 24.0 software^43^ was applied to statistically model the underlying mechanisms of the link between total sleep quality and DSS scores. In this analysis, a latent variable from brain GMV has been made and used in models. Mediation analysis investigates how much of the covariance between two variables can be explained by the mediator variable(s). Age, sex, and total GMV were controlled in mediation analyses as well. More details of mediation analysis are provided in the supplement.

## Results

### Demographics

The primary dataset of this investigation (HCP-Young) included 1101 participants (22–35 years, mean age = 28.79 ± 3.69, 54.3% female), 103 of whom (9%) had a history of DSM-IV-based depression episodes during their lifetime. The detailed demographic characteristics of participants are provided in **Table 1**. We had two other different datasets for out-of-cohort validation analysis i.e., the HCP-Aging and eNKI. We found 378 participants (36–59 years, mean age = 47.3 ± 7, 57.9% female) from the HCP-Aging dataset and 334 participants that had cross-sectional data (18–59 years, mean age = 37 ± 13.8, 62% female) from the eNKI dataset. From the eNKI dataset, we found 66 participants (20–56 years, mean age = 42 ± 9.7, 77.3% female) who had longitudinal records and there was one to five years gap between the two records across those individuals. Among them, there were 26 subjects (20– 45 years, mean age = 34 ± 8.2, 73.1% female) who received neurofeedback therapy between their first and second records and there was in average a 653-day gap between their records. While, the other 40 participants (36–56 years, mean age = 47 ± 6.1, 80% female), who had not received neurofeedback therapy, had an average of 847 days gap between their first and second visits.

**Table 1.**
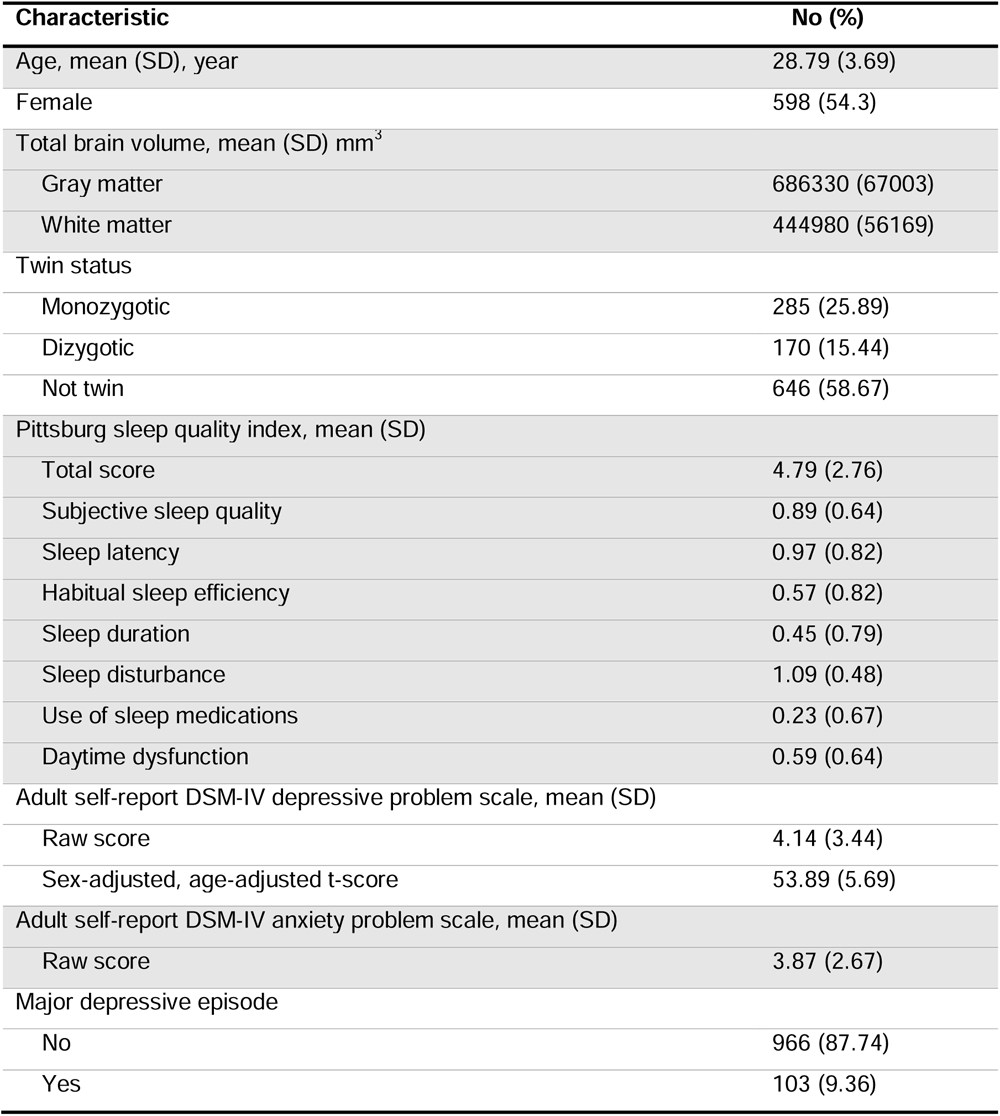
The demographic characteristics of 1101 participants from the HCP-Young dataset.

| Characteristic | No (%) |
| --- | --- |
| Age, mean (SD), year | 28.79 (3.69) |
| Female | 598 (54.3) |
| Total brain volume, mean (SD) mm<sup>3</sup> |  |
| Gray matter | 686330 (67003) |
| White matter | 444980 (56169) |
| Twin status |  |
| Monozygotic | 285 (25.89) |
| Dizygotic | 170 (15.44) |
| Not twin | 646 (58.67) |
| Pittsburg sleep quality index, mean (SD) |  |
| Total score | 4.79 (2.76) |
| Subjective sleep quality | 0.89 (0.64) |
| Sleep latency | 0.97 (0.82) |
| Habitual sleep efficiency | 0.57 (0.82) |
| Sleep duration | 0.45 (0.79) |
| Sleep disturbance | 1.09 (0.48) |
| Use of sleep medications | 0.23 (0.67) |
| Daytime dysfunction | 0.59 (0.64) |
| Adult self-report DSM-IV depressive problem scale, mean (SD) |  |
| Raw score | 4.14 (3.44) |
| Sex-adjusted, age-adjusted t-score | 53.89 (5.69) |
| Adult self-report DSM-IV anxiety problem scale, mean (SD) |  |
| Raw score | 3.87 (2.67) |
| Major depressive episode |  |
| No | 966 (87.74) |
| Yes | 103 (9.36) |

### Sleep and anxiety predicted DSS in the HCP-Young dataset

The details of ML pipeline for training and evaluation of models in the HCP-Young dataset are presented in **Fig. 1**. ML models based on sleep quality could predict DSS (unseen data during model training, r = 0.43, rMSE = 2.73, R^2^ = 0.18) (**Fig. 2A**). Adding anxiety score to sleep quality features improved the prediction drastically (r = 0.67, R^2^ = 0.45, rMSE = 2.25) (**Fig. 2B**). Whereas adding GMV features to the sleep quality (r = 0.41, R^2^ = 0.16, rMSE = 2.76) and combination of sleep quality and anxiety (r = 0.66, R^2^ = 0.44, rMSE = 2.26) did not improve their prediction (**Fig. 2C,D**). Based on the designed method, the ML algorithm automatically selected different feature numbers in each fold, but the selected hyperparameter of the method for all folds of all models was LS-boost.

Our complementary analyses demonstrated that brain morphological and functional features cannot predict DSS in general populations (**eFigure 2**). Removing participants with a history of depression showed robust predictive results e.g., a combination of sleep quality and anxiety predicted DSS similarly (r = 0.61, R^2^ = 0.37, rMSE = 2.18) (**eFigure 5**). Moreover, repeating the analyses based on seven components of PSQI (instead of 19 questions of the self-reported Pittsburgh sleep quality index (PSQI)) also revealed robust results in predicting DSS (r = 0.64, R^2^ = 0.41, rMSE = 2.32, based on a combination of sleep quality and anxiety) (**eFigure 6**). The feature importance in the ML model demonstrated that sleep-related daytime dysfunction, sleep disturbance, and subjective sleep quality were more important than other sleep components in predicting DSS (**eFigure 7**). Importantly, the reverse direction of prediction (prediction of sleep quality based on DSS) revealed a weaker result (r = 0.33, R^2^ = 0.11, rMSE = 2.61) (**eFigure 8**), indicating the sleep quality is a better predictor of DSS than another way around. Further, using the original DSS’ scores (not excluding two sleep-related questions from the depressive questionnaire) provides better prediction results (e.g. based on a combination of sleep quality and anxiety r = 0.71, R^2^ = 0.50, rMSE = 2.42) (**eFigure 10**). Then, we found that about 62% of the covariance between sleep quality and DSS’ is because of those two sleep-related items (**eFigure 11**). Moreover, we observed that 52.6% of the covariance between sleep quality and DSS is because of anxiety, while GMV could not mediate their association (**eFigure 12**). For more details, see the supplementary file.

### Sleep and anxiety predicted DSS in the independent datasets

Interestingly, we were able to predict DSS in both HCP-Aging and eNKI cohorts using models that were trained by the HCP-Young dataset (**Fig. 3A&B**). In the HCP-Aging dataset, sleep quality features could predict DSS robustly (r = 0.57, R^2^ = 0.27, rMSE = 2.64, CI = 3.27 – 3.54). Further, adding anxiety score to sleep quality features could improve the prediction in this dataset (r = 0.72, R^2^ = 0.50, rMSE = 2.19). Adding GMV features to the sleep quality (r = 0.56, R^2^ = 0.27, rMSE = 2.65) and a combination of sleep quality and anxiety score (r = 0.72, R^2^ = 0.49, rMSE = 2.21) provided similar results to the primary (HCP-Young) dataset.

Similarly, in the eNKI dataset, sleep quality predicted DSS (r = 0.50, R^2^ = 0.16, rMSE = 2.70), and a combination of sleep quality and anxiety score predicted DSS (r = 0.66, R^2^ = 0.38, rMSE = 2.34). Adding GMV features to the sleep quality could not improve the prediction (r = 0.51, R^2^ = 0.18, rMSE = 2.68), and a combination of sleep quality, anxiety, and GMV (r = 0.68, R^2^ = 0.40, rMSE = 2.29) revealed the same result as the HCP-Young dataset.

Finally, applying ML models on the longitudinal sample of the eNKI dataset resulted in the prediction of future depressive symptoms (**Fig. 4**) based on sleep quality (r = 0.61, R^2^ = 0.33, rMSE = 3.01) and combination of sleep quality and anxiety (r = 0.66, R^2^ = 0.44, rMSE = 2.73). Furthermore, results of confirmatory analyses in the longitudinal sample revealed that the predictability of DSS in subjects who had not received neurofeedback therapy between their first and second visits was strong (**eFigure 13A**). Interestingly, ML models were unable to predict future DSS when participants had received neurofeedback therapy between their first and second visits (**eFigure 13B**).

## Discussion

The main findings of this study highlighted that sleep quality could predict DSS in three independent datasets and adding anxiety to the sleep quality enhanced such prediction. (**Figure 2**). Structural and functional brain measurements neither predict DSS nor mediate the link between sleep quality and DSS. Our ML models provided similar results in other independent cohorts using cross-sectional (**Fig 3**), suggesting the generalizability of our ML models. Additionally, ML models’ performance was strong in the prediction of future individual DSS based on baseline sleep quality and anxiety in longitudinal samples (**Fig 4**). Our complementary analyses consider the impact of a lifetime history of depression, confounding effects of two sleep-related questions on the depressive assessment, the potential multicollinearity between variables, and lower predictive performance of the reverse direction (i.e., predicting sleep quality based on depressive symptoms) highlighted the robustness of our main findings. In addition, anxiety scores could mediate the association between sleep quality and DSS (**eFigure 12**).

Our findings are consistent with a body of literature showing that sleep disturbance and depressive problems are associated with each other in previous meta-analyses.^11–14^ In large-scale population cohorts, it has also been shown that sleep quality is associated with depressive symptoms ^9,25^. However, our study aimed to predict DSS based on sleep quality in different samples instead of investigating only the correlation between them. Animal models revealed that neonatal sleep disturbance could lead to adulthood depressive symptoms.^44,45^ Longitudinal human studies showed that people with sleep initiation problems might experience depression over the next 3-6 years of their life.^46,47^ Interestingly, toddlers’ sleep problems at the age of 18 months predicted depressive symptoms at the age of 8 years old.^48^ Several large-scale longitudinal studies^49–52^ demonstrated that short sleep duration and sleep disturbance should be considered risk factors for developing future depressive symptoms. Although these studies have not used ML models to be able to predict individual DSS within the same sample or other samples, they suggest that poor sleep could be a critical predictor for DSS. A recent ML study ^53^ demonstrated that sleep disorder is one of the most important features to predict depression, particularly in individuals with hypertension. They predicted a binary definition (existence/nonexistence) of depression among adults with hypertension, while our study predicted a wide (0-28) continuous range of severity of depressive symptoms in three databases. Another large-scale ML-based study found that sleep duration is one of the top five predictors of DSS among home-based older adults.^54^ Our findings support this hypothesis, although they do not claim to show any causality between sleep and DSS. The cross-sectional nature of our study precludes the assessment of the long-term causal pathways in the general population. Thus, the longitudinal role of poor sleep (using both subjective and objective sleep assessments) in developing clinical MDD has to be examined in the future.

In the present study, anxiety improved the prediction of DSS by sleep quality features (**Fig 2**). As it is described in **eFigure 3**, although anxiety alone could predict DSS (r = 0.62), the correlation coefficient between anxiety and DSS itself was already robust (r = 0.63) (**eFigure 11**), which means the ML model based on anxiety features (alone) was not superior. However, while the baseline correlation between sleep quality and DSS was r = 0.24 (**eFigure 11**), ML models could predict DSS based on sleep quality with r = 0.34 (**Fig 2**) which shows the better performance of the ML model based on sleep quality. So, we can suggest that sleep quality is the predictor of DSS and anxiety strengths it’s prediction. Anxiety scores had also an indirect effect (51.2%) in mediating the link between sleep quality and DSS. The strong interplay between sleep disturbances, anxiety, and depression has been well-documented earlier,^23,55^ and our study supports such findings. For example, short and long sleep duration are predictors of depression and anxiety in a large cohort.^56^ The additive role of anxiety to sleep in DSS prediction is further supported by the notion that sleep loss increases preemptive responding in the amygdala and anterior insula during affective anticipation.^57^

Previous studies have shown that poor sleep loss is linked to abnormal activity in the medial prefrontal cortex, amygdala, insula, and anterior cingulate cortex, which were associated with higher levels of next-day anxiety.^58^ An earlier study using the HCP-Young sample indicated that functional connectivity between the lateral orbitofrontal cortex, dorsolateral prefrontal cortex, anterior and posterior cingulate cortices, insula, parahippocampal gyrus, hippocampus, amygdala, temporal cortex, and precuneus mediated the effect of sleep quality on DSS.^25^ Structural brain alterations in the postcentral gyrus and superior temporal gyrus mediate the link between sleep disturbance and depressive symptoms in a small group of shift-working nurses.^59^ Other studies observed that the GMV of the right insula mediates the relationship between sleep quality and anxiety/depressive symptoms among young students.^26,60^ However, these studies have mainly assessed the association between sleep quality and depressive symptoms and the brain and have not focused on inter-individual prediction. In the present study, GMV did not predict DSS in any dataset, could not improve prediction performance when combined with sleep and anxiety features, and did not have any mediatory effect on the link between sleep and DSS. One explanation could be the link between sleep disturbance and depressive symptoms anchored in the functional level rather than GMV.^24^ However, our complementary analyses showed that ReHo and fALFF features of brain function could not predict DSS as well. Although a previous study found that functional connectivity across the brain is a better predictor for behavioral measures than anatomical and diffusion features, they did not assess the predictability of depressive problems.^61^ In this study, the brain measurements were associated with sleep quality (**eFigure 11**) but were not predictors or mediators of DSS and were not correlated with DSS, which according to the amount of sleep quality (mean = 4.79, SD = 2.76 the average of total scores is close to the threshold of poor sleep quality 5) and DSS (mean _t-score_ = 53.89, SD _t-score_ = 5.69 the average of t-scores is far less than clinical range 69) in the HCP-Young dataset (**Table 1**), in average the level of poor sleep quality and DSS in this dataset was not so prominent to be appearing in the brain. Another important point of this study is that we excluded two sleep-related items from the DSS questionnaire. As it is shown in **eFigure 11**, when we included sleep-related items, the association between sleep quality and DSS score increased, and we found some brain areas correlated with DSS scores (similar to previous studies) which were mainly due to those sleep-related items of depressive problems questionnaire. The brain-related results of our study were similar to previous large-scale neuroimaging meta-analyses studies which failed to identify a robust regional abnormality in clinical insomnia disorder, MDD, and late-life depression.^62–64^ Similarly, the ML classification model failed to separate healthy individuals from subjects with insomnia based on brain volumes ^65^ or to differentiate healthy individuals from patients with depression based on brain structure and function values,^66^ indicating that the neurobiological underpinning mechanism of sleep disorders and depression is still under debate and needs further elaboration.

In conclusion, we found that sleep quality could predict DSS across cross-sectional and longitudinal samples. Anxiety symptoms, rather than brain features, improved the performance of the predictive model and mediated the link between sleep and DSS. Although the sample size of our samples (mainly for our longitudinal analyses) was small, our ML models have shown the generalizability of their outcomes in different cohorts. Future large-scale longitudinal datasets are needed to assess the role of sleep and anxiety on the development of depressive symptoms and clinical MDD in the general population. We hope that our findings incentivize clinicians to consider the importance of screening and treating subjects with sleep disturbance and anxiety problems to reduce the burden of depressive symptoms in the general population.

## Author Contributions

Conception and study design: M.O., S.B.E. and M.T. Preprocessing and data analysis: M.O., F.S., S.F., S.M., K.P. Interpretation M.O., S.G., S.B.E., and M.T. Paper writing and editing: all authors.

## Conflict of Interest Disclosures

The authors declare no conflicts of interest.

## Role of the funding source

The funders had no role in the design and conduct of the study; collection, management, analysis, and interpretation of the data; preparation, review, or approval of the manuscript; and the decision to submit the manuscript for publication.

## Data Sharing Statement

We used the public datasets from the Human Connectome Project (https://www.humanconnectome.org/) and the enhanced Nathan Kline Institute-Rockland sample (eNKI) (http://fcon_1000.projects.nitrc.org/indi/enhanced/).

## Data Availability

All data produced in the present study are available upon reasonable request to the authors

## Notes

### Competing Interest Statement

The authors have declared no competing interest.

### Funding Statement

This study did not receive any funding

### Summary of Updates

Several new complementary analyses have been added to this version.

## References

1. Otte C, Gold SM, Penninx BW, et al. Major depressive disorder. Nature Reviews Disease Primers 2016; 2(1): 16065.

2. Wang J, Wu X, Lai W, et al. Prevalence of depression and depressive symptoms among outpatients: a systematic review and meta-analysis. BMJ Open 2017; 7(8): e017173.

3. Keyes KM, Gary D, O’Malley PM, Hamilton A, Schulenberg J. Recent increases in depressive symptoms among US adolescents: trends from 1991 to 2018. Social Psychiatry and Psychiatric Epidemiology 2019; 54(8): 987–96.

4. Ettman CK, Abdalla SM, Cohen GH, Sampson L, Vivier PM, Galea S. Prevalence of Depression Symptoms in US Adults Before and During the COVID-19 Pandemic. JAMA Network Open 2020; 3(9): e2019686-e.

5. Moazen-Zadeh E, Assari S. Depressive Symptoms Predict Major Depressive Disorder after 15 Years among Whites but Not Blacks. Front Public Health 2016; 4: 13.

6. Cuijpers P, Reynolds CF, III. Increasing the Impact of Prevention of Depression—New Opportunities. JAMA Psychiatry 2022; 79(1): 11–2.

7. Greenberg PE, Fournier AA, Sisitsky T, et al. The Economic Burden of Adults with Major Depressive Disorder in the United States (2010 and 2018). Pharmacoeconomics 2021; 39(6): 653–65.

8. Becker NB, Jesus SN, Joao K, Viseu JN, Martins RIS. Depression and sleep quality in older adults: a meta-analysis. Psychol Health Med 2017; 22(8): 889–95.

9. Huang Y, Zhu M. Increased Global PSQI Score Is Associated with Depressive Symptoms in an Adult Population from the United States. Nature and Science of Sleep 2020; 12: 487–95.

10. Riemann D, Krone LB, Wulff K, Nissen C. Sleep, insomnia, and depression. Neuropsychopharmacology 2020; 45(1): 74–89.

11. Baglioni C, Battagliese G, Feige B, et al. Insomnia as a predictor of depression: a meta-analytic evaluation of longitudinal epidemiological studies. J Affect Disord 2011; 135(1-3): 10–9.

12. Hertenstein E, Feige B, Gmeiner T, et al. Insomnia as a predictor of mental disorders: A systematic review and meta-analysis. Sleep Medicine Reviews 2019; 43: 96–105.

13. Li L, Wu C, Gan Y, Qu X, Lu Z. Insomnia and the risk of depression: a meta-analysis of prospective cohort studies. BMC Psychiatry 2016; 16(1): 375.

14. Emamian F, Khazaie H, Okun ML, Tahmasian M, Sepehry AA. Link between insomnia and perinatal depressive symptoms: A meta-analysis. J Sleep Res 2019; 28(6): e12858.

15. Franzen PL, Buysse DJ. Sleep disturbances and depression: risk relationships for subsequent depression and therapeutic implications. Dialogues in clinical neuroscience 2008; 10(4): 473–81.

16. Khazaie H, Ghadami MR, Knight DC, Emamian F, Tahmasian M. Insomnia treatment in the third trimester of pregnancy reduces postpartum depression symptoms: a randomized clinical trial. Psychiatry Res 2013; 210(3): 901–5.

17. Irwin MR, Carrillo C, Sadeghi N, Bjurstrom MF, Breen EC, Olmstead R. Prevention of Incident and Recurrent Major Depression in Older Adults With Insomnia: A Randomized Clinical Trial. JAMA Psychiatry 2022; 79(1): 33–41.

18. Wassing R, Benjamins JS, Talamini LM, Schalkwijk F, Van Someren EJW. Overnight worsening of emotional distress indicates maladaptive sleep in insomnia. Sleep 2019; 42(4).

19. Meneo D, Samea F, Tahmasian M, Baglioni C. The emotional component of insomnia disorder: A focus on emotion regulation and affect dynamics in relation to sleep quality and insomnia. Journal of Sleep Research 2023; 32(6): e13983.

20. Wassing R, Benjamins JS, Dekker K, et al. Slow dissolving of emotional distress contributes to hyperarousal. Proc Natl Acad Sci U S A 2016; 113(9): 2538–43.

21. Schiel JE, Holub F, Petri R, et al. Affect and Arousal in Insomnia: Through a Lens of Neuroimaging Studies. Current Psychiatry Reports 2020; 22(9): 44.

22. Ebneabbasi A, Mahdipour, M., Nejati, V., Li, M., Liebe, T., Colic, L., Leutritz, A.L., Vogel, M., Zarei, M., Walter, M., Tahmasian, M. Emotion processing and regulation in major depressive disorder: A 7T resting-state fMRI stud. PsyArXiv 2020.

23. Alvaro PK, Roberts RM, Harris JK. A systematic review assessing bidirectionality between sleep disturbances, anxiety, and depression. Sleep 2013; 36(7): 1059–68.

24. Bagherzadeh-Azbari S, Khazaie H, Zarei M, et al. Neuroimaging insights into the link between depression and Insomnia: A systematic review. Journal of Affective Disorders 2019; 258: 133–43.

25. Cheng W, Rolls ET, Ruan H, Feng J. Functional Connectivities in the Brain That Mediate the Association Between Depressive Problems and Sleep QualityFunctional Connectivities in the Brain That Mediate the Association Between Depression and Sleep QualityFunctional Connectivities in the Brain That Mediate the Association Between Depression and Sleep Quality. JAMA Psychiatry 2018; 75(10): 1052–61.

26. Yulin Wang YT, Zhiliang Long, Debo Dong, Qinghua He Jiang Qiu,, Tingyong Feng HC, Masoud Tahmasian, Xu Lei. The volume of the Dentate Gyrus/CA4 Hippocampal Subfield Mediates the Interplay between Sleep Quality and Depressive Symptoms. 2023.

27. Goldstein-Piekarski AN, Holt-Gosselin B, O’Hora K, Williams LM. Integrating sleep, neuroimaging, and computational approaches for precision psychiatry. Neuropsychopharmacology 2020; 45(1): 192–204.

28. Williams LM. Precision psychiatry: a neural circuit taxonomy for depression and anxiety. The Lancet Psychiatry 2016; 3(5): 472–80.

29. Van Essen DC, Ugurbil K, Auerbach E, et al. The Human Connectome Project: A data acquisition perspective. NeuroImage 2012; 62(4): 2222–31.

30. 10.15154/1520707.

31. Bookheimer SY, Salat DH, Terpstra M, et al. The Lifespan Human Connectome Project in Aging: An overview. Neuroimage 2019; 185: 335–48.

32. Nooner KB, Colcombe SJ, Tobe RH, et al. The NKI-Rockland Sample: A Model for Accelerating the Pace of Discovery Science in Psychiatry. Front Neurosci 2012; 6: 152.

33. Achenbach System of Empirically Based Assessment (ASEBA). Adult (Ages 18-59) Assessments.. https://aseba.org/adults/.

34. Glasser MF, Sotiropoulos SN, Wilson JA, et al. The minimal preprocessing pipelines for the Human Connectome Project. NeuroImage 2013; 80: 105–24.

35. Christian Gaser RD. CAT-A Computational Anatomy Toolbox for the Analysis of Structural MRI Data. 22nd Annual Meeting of the Organization for Human Brain Mapping 2016.

36. Schaefer A, Kong R, Gordon EM, et al. Local-Global Parcellation of the Human Cerebral Cortex from Intrinsic Functional Connectivity MRI. Cereb Cortex 2018; 28(9): 3095–114.

37. Fan L, Li H, Zhuo J, et al. The Human Brainnetome Atlas: A New Brain Atlas Based on Connectional Architecture. Cereb Cortex 2016; 26(8): 3508–26.

38. Yeo BT, Krienen FM, Sepulcre J, et al. The organization of the human cerebral cortex estimated by intrinsic functional connectivity. J Neurophysiol 2011; 106(3): 1125–65.

39. Mohajer B, Abbasi N, Mohammadi E, et al. Gray matter volume and estimated brain age gap are not linked with sleep-disordered breathing. Hum Brain Mapp 2020; 41(11): 3034–44.

40. MATLAB (2020) Version 9.8.0.1417392 (R2020a). The MathWorks Inc., Natick.

41. Robnik-Sikonja M, Kononenko I. An adaptation of Relief for attribute estimation in regression. ICML; 1997; 1997.

42. Practical Bayesian Optimization of Machine Learning Algorithms. 2012.

43. Arbuckle JL. Amos (Version 24.0) [Computer Program]. *Chicago: SPSS* 2019.

44. Savelyev SA, Rantamaki T, Rytkonen KM, Castren E, Porkka-Heiskanen T. Sleep homeostasis and depression: studies with the rat clomipramine model of depression. Neuroscience 2012; 212: 149–58.

45. Mendoza J. Circadian insights into the biology of depression: Symptoms, treatments and animal models. Behavioural Brain Research 2019; 376: 112186.

46. Blanken TF, Borsboom D, Penninx BW, Van Someren EJ. Network outcome analysis identifies difficulty initiating sleep as a primary target for prevention of depression: a 6-year prospective study. Sleep 2019; 43(5).

47. Riemann D, Voderholzer U. Primary insomnia: a risk factor to develop depression? Journal of Affective Disorders 2003; 76(1): 255–9.

48. Sivertsen B, Harvey AG, Reichborn-Kjennerud T, Ystrom E, Hysing M. Sleep problems and depressive symptoms in toddlers and 8-year-old children: A longitudinal study. J Sleep Res 2021; 30(1): e13150.

49. Li Y, Wu Y, Zhai L, Wang T, Sun Y, Zhang D. Longitudinal Association of Sleep Duration with Depressive Symptoms among Middle-aged and Older Chinese. Scientific Reports 2017; 7(1): 11794.

50. Marino C, Andrade B, Montplaisir J, et al. Testing Bidirectional, Longitudinal Associations Between Disturbed Sleep and Depressive Symptoms in Children and Adolescents Using Cross-Lagged Models. JAMA Network Open 2022; 5(8): e2227119-e.

51. Suh S, Kim H, Yang H-C, Cho ER, Lee SK, Shin C. Longitudinal Course of Depression Scores with and without Insomnia in Non-Depressed Individuals: A 6-Year Follow-Up Longitudinal Study in a Korean Cohort. Sleep 2013; 36(3): 369–76.

52. Yokoyama E, Kaneita Y, Saito Y, et al. Association between Depression and Insomnia Subtypes: A Longitudinal Study on the Elderly in Japan. Sleep 2010; 33(12): 1693–702.

53. Lee C, Kim H. Machine learning-based predictive modeling of depression in hypertensive populations. PLoS One 2022; 17(7): e0272330.

54. Lin S, Wu Y, He L, Fang Y. Prediction of depressive symptoms onset and long-term trajectories in home-based older adults using machine learning techniques. Aging Ment Health 2022: 1–10.

55. Oh C-M, Kim HY, Na HK, Cho KH, Chu MK. The Effect of Anxiety and Depression on Sleep Quality of Individuals With High Risk for Insomnia: A Population-Based Study. Frontiers in Neurology 2019; 10.

56. van Mill JG, Vogelzangs N, van Someren EJ, Hoogendijk WJ, Penninx BW. Sleep duration, but not insomnia, predicts the 2-year course of depressive and anxiety disorders. J Clin Psychiatry 2014; 75(2): 119–26.

57. Goldstein AN, Greer SM, Saletin JM, Harvey AG, Nitschke JB, Walker MP. Tired and Apprehensive: Anxiety Amplifies the Impact of Sleep Loss on Aversive Brain Anticipation. The Journal of Neuroscience 2013; 33(26): 10607–15.

58. Ben Simon E, Rossi A, Harvey AG, Walker MP. Overanxious and underslept. Nature Human Behaviour 2020; 4(1): 100–10.

59. Park CH, Bang M, Ahn KJ, Kim WJ, Shin NY. Sleep disturbance-related depressive symptom and brain volume reduction in shift-working nurses. Sci Rep 2020; 10(1): 9100.

60. Yin H, Zhang L, Li D, Xiao L, Cheng M. The gray matter volume of the right insula mediates the relationship between symptoms of depression/anxiety and sleep quality among college students. J Health Psychol 2019: 1359105319869977.

61. Ooi LQR, Chen J, Zhang S, et al. Comparison of individualized behavioral predictions across anatomical, diffusion and functional connectivity MRI. Neuroimage 2022; 263: 119636.

62. Tahmasian M, Noori K, Samea F, et al. A lack of consistent brain alterations in insomnia disorder: An activation likelihood estimation meta-analysis. Sleep Med Rev 2018; 42: 111–8.

63. Muller VI, Cieslik EC, Serbanescu I, Laird AR, Fox PT, Eickhoff SB. Altered Brain Activity in Unipolar Depression Revisited: Meta-analyses of Neuroimaging Studies. JAMA Psychiatry 2017; 74(1): 47–55.

64. Saberi A, Mohammadi E, Zarei M, Eickhoff SB, Tahmasian M. Structural and functional neuroimaging of late-life depression: a coordinate-based meta-analysis. Brain Imaging Behav 2022; 16(1): 518–31.

65. Weihs A, Frenzel S, Bi H, et al. Lack of structural brain alterations associated with insomnia: findings from the ENIGMA-Sleep Working Group. J Sleep Res 2023: e13884.

66. Winter NR, Leenings R, Ernsting J, et al. Quantifying Deviations of Brain Structure and Function in Major Depressive Disorder Across Neuroimaging Modalities. JAMA Psychiatry 2022; 79(9): 879–88.

