## Supplemental file for "Prediction of depressive symptoms severity based on sleep quality, anxiety, and brain: a machine learning approach across three cohorts"

**eMethod**

**eFigure 1.** Correlation between sleep quality features.

**eTable 1.** Sleep quality assessment using PSQI.

**eFigure 2.** Prediction of DSS in the HCP-Young dataset based on neuroimaging features (GMV, ReHo, fALFF).

**eFigure 3.** Prediction of DSS based on anxiety and combination of GMV and anxiety.

**eFigure 4.** Cross-prediction of DSS and anxiety.

**eFigure 5.** Prediction of DSS after excluding participants who experienced depression.

**eFigure 6.** Prediction of DSS based on seven components of PSQI.

**eFigure 7.** Feature importance of 7 PSQI components.

**eFigure 8.** Prediction of sleep quality based on DSS.

**eFigure 9.** Prediction of sleep quality based on GMV.

**eFigure 10.** Prediction of DSS’ in the HCP-Young dataset after including two sleep-related items of depressive symptoms severity.

**eFigure 11.** Comparison of DSS and DSS’.

**eFigure 12.** Mediation analysis of GMV and anxiety on the link between sleep quality and DSS.

**eFigure 13.** Longitudinal prediction in the eNKI sample after separating subjects who had received neurofeedback therapy.

**Participants and data preprocessing**

This investigation had 1101 subjects from the HCP-Young dataset for main analyses, 334 subjects from the eNKI, and 378 subjects from the HCP-Aging datasets for out-of-cohort validation of trained ML models, obtained from the HCP-Young. All participants who had phenotypic and MRI neuroimaging data were included and this was the inclusion criteria in this study. The phenotypical data (sleep quality, anxiety, DSS) were measured by the same questionnaires in all three datasets. In addition, the HCP-Young dataset has provided information about the history of diagnosed depression with the question “Has the participant experienced a diagnosed DSM-IV major depressive episode over his/her lifetime?”. We excluded participants who had at least one episode of depression in confirmatory analyses (**eFigure 3**) to test the robustness of models for participants who never had a history of depression. In addition, we used structural MRI images of the HCP-Young dataset, which were acquired by Siemens 3T Skyra scanner and preprocessed using the HCP pipelines.^1^ These T1 weighted MRI images were collected with 0.7mm voxel size isotropic resolution, time of repetition (TR) = 2400 ms, time of echo (TE) = 2.14 ms, time of inversion (TI) = 1000 ms, with flip angle of 8 degrees, and the field of view was 224×224 mm.^2^

**Calculation of local measures of resting-state functional MRI**

Local measures of intrinsic brain activity were assessed as regional homogeneity (ReHo) and fractional amplitude of low-frequency fluctuations (fALFF) using 3T whole-brain resting-state functional MRI (rsfMRI) data with 2mm isotropic spatial resolution and a repetition time of 0.72 seconds.

We used minimally preprocessed data from all four rsfMRI recordings provided by the HCP. During the preprocessing, the image underwent gradient distortion correction, motion correction, image distortion correction, registration to the T1 structural MRI image, normalization to standard space MNI, normalization to the intensity of the rsfMRI image global mean, and artifact removal using ICA-FIX.^1,3^ Additional steps, conducted using the Junifer toolbox,^4^ based on AFNI functions,^5^ included regressing out the white matter, cerebrospinal fluid, global signal, and motion parameters, followed by linearly detrending and bandpass filtering (0.01-0.08 Hz for ReHo, 0.01-0.1 Hz for fALFF) within a grey-matter mask.

ReHo, reflecting the similarity of each voxel’s time series with its nearest neighbors was computed using Kendall’s coefficient of concordance. This measures the similarity of time series between neighboring voxels.^6^ fALFF indicates the ratio of power within the 0.01-0.1 Hz frequency range to the total spectral power without bandpass filtering. Hereby, each voxel’s time series is transformed to the frequency domain, the square root of the power spectrum at each frequency and summing the power within the frequency range divided by the total power spectrum per voxel.^7^ Both ReHo and fALFF were calculated parcel-wise across a combination of 400 cortical parcels from Schaefer atlas,^8^ 36 subcortical parcels from Brainnetome,^9^ and 37 cerebellar parcels from Buckner,^10^ as previously discussed for the VBM analysis. Here all voxel-wise values in each parcel were averaged. We further averaged the parcel-wise scores across available recordings for each participant. Participants with QC issue code B: Segmentation and Surface QC were excluded.

**Machine learning-based prediction**

**ML models**

The ensemble decision tree model was used in this investigation, which is one of the most interpretable and powerful ML techniques available. Hyperparameters of these models were the ensemble aggregation method (bagging/LS-Boost), number of ensemble learning cycles [10,50], learning rate (0,1], and minimum number of leaf node observations. Noticeably, in all models, LS-Boost was selected as the best method in the optimization process. ML pipeline in this investigation consisted of three sequential steps (cross-validation, feature selection, model training, and evaluation):

**Cross-validation:** First of all, a standard nested 10-fold cross-validation was performed to assess the generalizability of models. The following protocol was applied for nested 10-fold cross-validation: HCP subjects were divided into 10 non-overlapping folds. Each fold was used as a held-back test set, whilst all other folds collectively constructed the training set. Another 10-fold cross-validation was applied to each training set and made 10 validation sets in each training set. Further, to consider the family structure of HCP subjects in ML analyses, we paid special attention that siblings were not separated in train/validation/test sets (**Fig 2**).

**Feature selection:** In models with 473 GMV features (**Fig 3B&D**), a filter-based feature selection method was applied to reduce the computational cost, prevent overfitting, and improve model performance. Hence, features of training sets were ranked and scored by the relief method (If there is a feature value difference in a neighboring instance pair with a different target, the relief method increases the feature rank ^11^). Afterward, 10 different numbers of top-scoring features (20, 30, 40, 50, 60, 70, 80, 90, 100, 110 highest-scoring features) were checked in each inner fold for model training so the algorithm could select the best feature number in each fold automatically. However, in models with only 19 PSQI or 20 PSQI and anxiety features (**Fig 3A&C**), we did not perform feature selection because there was no feature redundancy (as we checked the non-existence of feature redundancy and collinearity between sleep quality features by correlation analysis **eFigure 2**.) and the number of features was not so high.

**Model training and evaluation:** Model training and hyperparameter optimization with 100 iterations were performed in the inner loop of cross-validation and repeated 10 times with 10 different feature numbers. Therefore, a total of 1000 models (10 outer loop * 10 inner loop * 10 times with different feature numbers) and 100000 iterations (1000 model * 100 iterations) were evaluated by validation sets, and then 10 models with the minimum MSE were selected. These models were fitted on the entire training sets and evaluated by the unseen test sets. Finally, the average performance of models is reported in **Fig .3**.

**Complementary analyses**

We performed several follow-up analyses in the HCP-Young dataset to show the robustness of our results. First of all, **eFigure 1** shows the correlation between sleep quality features in which we didn’t perform feature selection because there is no redundancy nor collinearity between these features based on their correlation. The correlation coefficient between total sleep quality and DSS (r = 0.29 p-value < 0.001), and total anxiety (r = 0.32 p-value < 0.001) and also between DSS and anxiety (r = 0.63, p-value < 0.001) were strong and there was no significant correlation between GMV/ReHo/fALFF and DSS/anxiety. Then we assessed the predictability of individualized variability of DSS based on brain structural and functional parameters (**eFigure 2**). Then, we predicted DSS based on only anxiety scores and a combination of anxiety and GMV features to test the predictability of anxiety itself and in combination with GMV (**eFigure 3**). At the next stage, we performed cross-prediction of DSS and anxiety to test collinearity between them (**eFigure 4**). For the next confirmatory analysis, we excluded participants who had experienced at least one episode of depression to test the robustness of our results (**eFigure 5**) in healthy people without any history of depression. Further, we predicted DSS based on seven components of PSQI (**eFigure 6**) and assessed the most important features in this model (**eFigure 7**). Moreover, we assessed the reverse direction of prediction by predicting sleep quality based on DSS (**eFigure 8**) and GMV (**eFigure 9**). Then, we included sleep-related scores of depressive problems measure (named these scores as DSS’) and compared the results before and after their inclusion (**eFigure 10&11**). Finally, we tested the mechanisms underlying the link between sleep quality and DSS (**eFigure 12**). As the last confirmatory analysis, we assessed the predictability of DSS in the longitudinal sample of the eNKI separating participants who had or had not received neurofeedback therapy (**eFigure 13**).

**Mediation analyses**

Standard mediation analyses were performed using Amos v.24 software, ^12^ which is a powerful software in path analysis and structural equation modeling (SEM). We performed mediation analysis using SEM to statistically model the underlying mechanisms of the link between total sleep quality and DSS scores. Mediation analysis investigates how much of the covariance between two variables can be explained by the mediator variable(s).

The main hypothesis of the mediation analysis was to assess the underlying mechanisms of the association between sleep quality and DSS. Hence, we made a latent variable of brain GMVs that had a significant correlation with sleep quality and used it in the mediation model as the GMV. Of note, there was no GMV parcel correlated with DSS or anxiety. So, the mediational role of anxiety and GMV were assessed in parallel in a single mediation model **eFigure 12.** In this analysis, age, sex, and total GMV were controlled and the significance of the models was tested by bias-corrected bootstrapping using 5000 random subsampling.

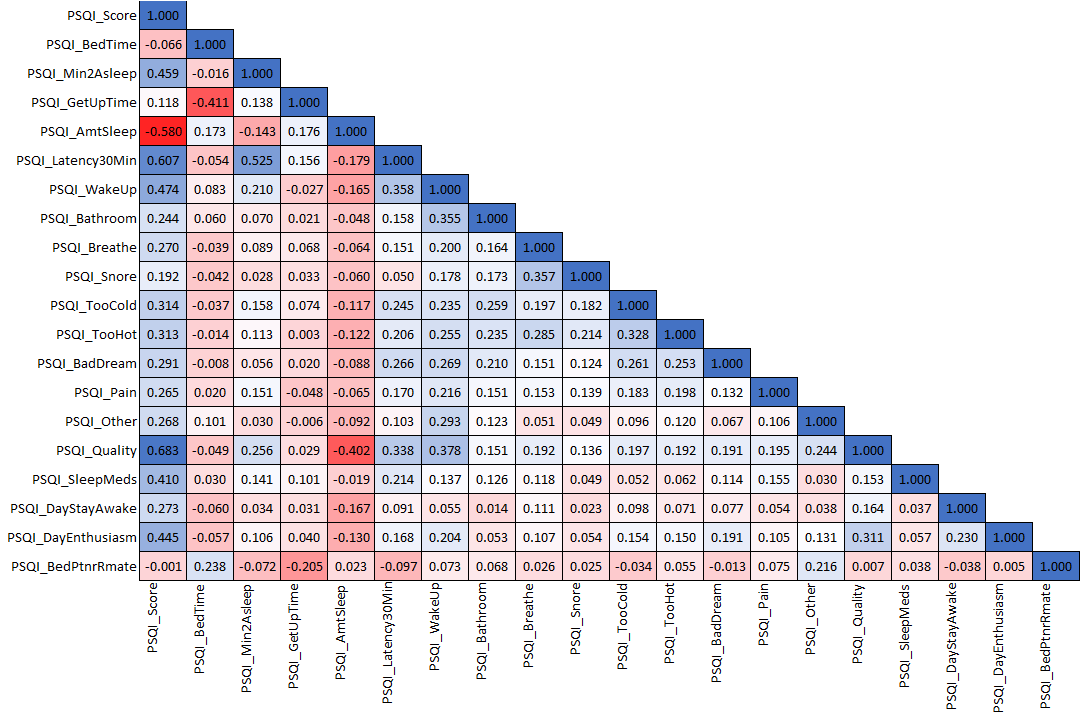

**eFigure 2.** Correlation between sleep quality features. All correlations are significant p-value < 0.05 (PSQI: Pittsburgh Sleep Quality Index, For more details, see **eTable 1**).

**eTable 1. Sleep quality assessment using PSQI**

| PSQI_Score | The total score across all items on the PSQI ^13^. |
| --- | --- |
| PSQI_BedTime | During the past month, when have you usually gone to bed at night? |
| PSQI_Min2Asleep | During the past month, how long has it usually taken you to fall asleep each night. |
| PSQI_GetUpTime | During the past month, when have you usually gotten up in the morning? |
| PSQI_AmtSleep | During the past month, how many hours of actual sleep did you get at night? (This may be different than the number of hours you spend in bed.) |
| PSQI_Latency30Min | During the past month, how often have you had trouble sleeping because you... (a) Cannot get to sleep within 30 minutes; 0=Not during the past month, 1=Less than once a week, 2=Once or twice a week, 3=3 or more times a week |
| PSQI_WakeUp | During the past month, how often have you had trouble sleeping because you... (b) Wake up in the middle of the night or early morning; 0=Not during the past month, 1=Less than once a week, 2=Once or twice a week, 3=3 or more times a week |
| PSQI_Bathroom | During the past month, how often have you had trouble sleeping because you... I Have to get up to use the bathroom; 0=Not during the past month, 1=Less than once a week, 2=Once or twice a week, 3=3 or more times a week |
| PSQI_Breathe | During the past month, how often have you had trouble sleeping because I... (d) Cannot breathe comfortably; 0=Not during the past month, 1=Less than once a week, 2=Once or twice a week, 3=3 or more times a week |
| PSQI_Snore | During the past month, how often have you had trouble sleeping becauIu... (e) Cough or snore loudly; 0=Not during the past month, 1=Less than once a week, 2=Once or twice a week, 3=3 or more times a week |
| PSQI_TooCold | During the past month, how often have you had trouble sleeping Iause you... (f) Feel too cold; 0=Not during the past month, 1=Less than once a week, 2=Once or twice a week, 3=3 or more times a week |
| PSQI_TooHot | During the past month, how often have you had trouble sleepiIbecause you... (g) Feel too hot; 0=Not during the past month, 1=Less than once a week, 2=Once or twice a week, 3=3 or more times a week |
| PSQI_BadDream | During the past month, how often have you had trouble sleIng because you... (h) Had bad dreams; 0=Not during the past month, 1=Less than once a week, 2=Once or twice a week, 3=3 or more times a week |
| PSQI_Pain | During the past month, how often have you had trouble Ieping because you... (i) Have pain; 0=Not during the past month, 1=Less than once a week, 2=Once or twice a week, 3=3 or more times a week |
| PSQI_Other | During the past month, how often have you had trouI sleeping because of... (j) Other reason(s), as described in 5j. pt2 0=Not during the past month, 1=Less than once a week, 2=Once or twice a week, 3=3 or more times a week |
| PSQI_Quality | During the past month, how would you rate your sleep quality overall? 0=Very good, 1=Fairly good, 2=Fairly bad, 3=Very bad |
| PSQI_SleepMeds | During the past month, how often have you taken”medicine (prescri”ed or \"over the counter\") to help you sleep? 0=Not during the past month, 1=Less than once a week, 2=Once or twice a week, 3=3 or more times a week |
| PSQI_DayStayAwake | During the past month, how often have you had trouble staying awake while driving, eating meals, or engaging in social activity? 0=Not during the past month, 1=Less than once a week, 2=Once or twice a week, 3=3 or more times a week |
| PSQI_DayEnthusiasm | During the past month, how much of a problem has it been for you to keep up enough enthusiasm to get things done? 0=No problem at all, 1=Only a very slight problem, 2=Somewhat of a problem, 3=A very big problem |
| PSQI_BedPtnrRmate | Do you have a bed partner or roommate? 0=No bed partner or roommate, 1=Partner/roommate in other room, 2=Partner in same room, but not same bed, 3=Partner in same bed |

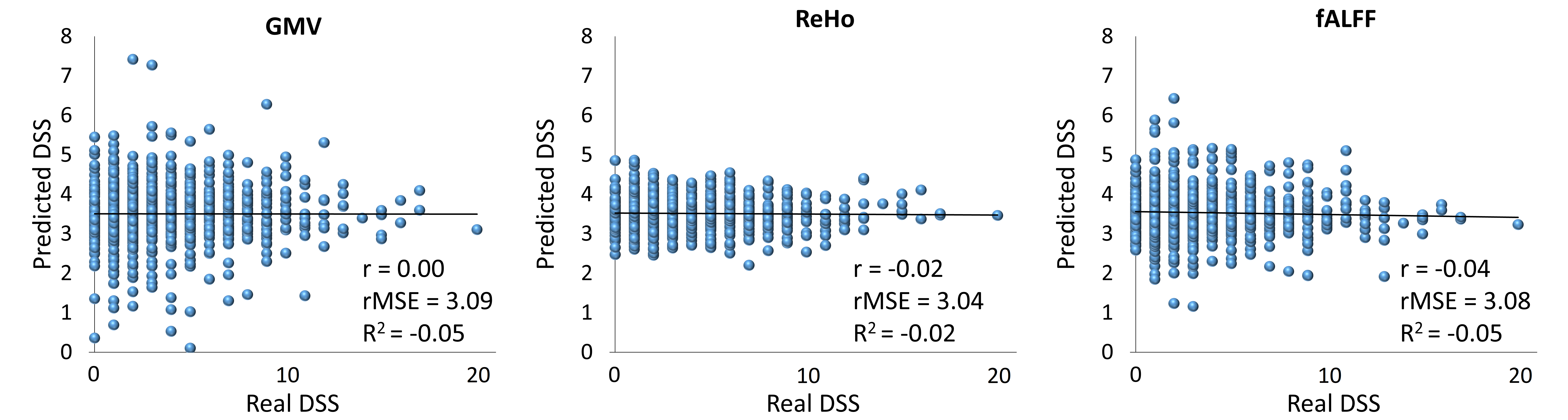

**eFigure 2.** Prediction of DSS in HCP-Young dataset based on neuroimaging features (GMV, ReHo, fALFF). (DSS: depressive symptoms severity after excluding two sleep-related items, GMV: gray matter volume, ReHo: regional homogeneity, fALFF: fractional amplitude of low-frequency fluctuations, r: correlation coefficient between real and predicted DSS, rMSE: root mean squared error, R^2^: determination coefficient).

The results of DSS prediction based on neuroimaging data show that we cannot predict the severity of depressive symptoms in general populatio’s based on only the brain's morphological and functional features.

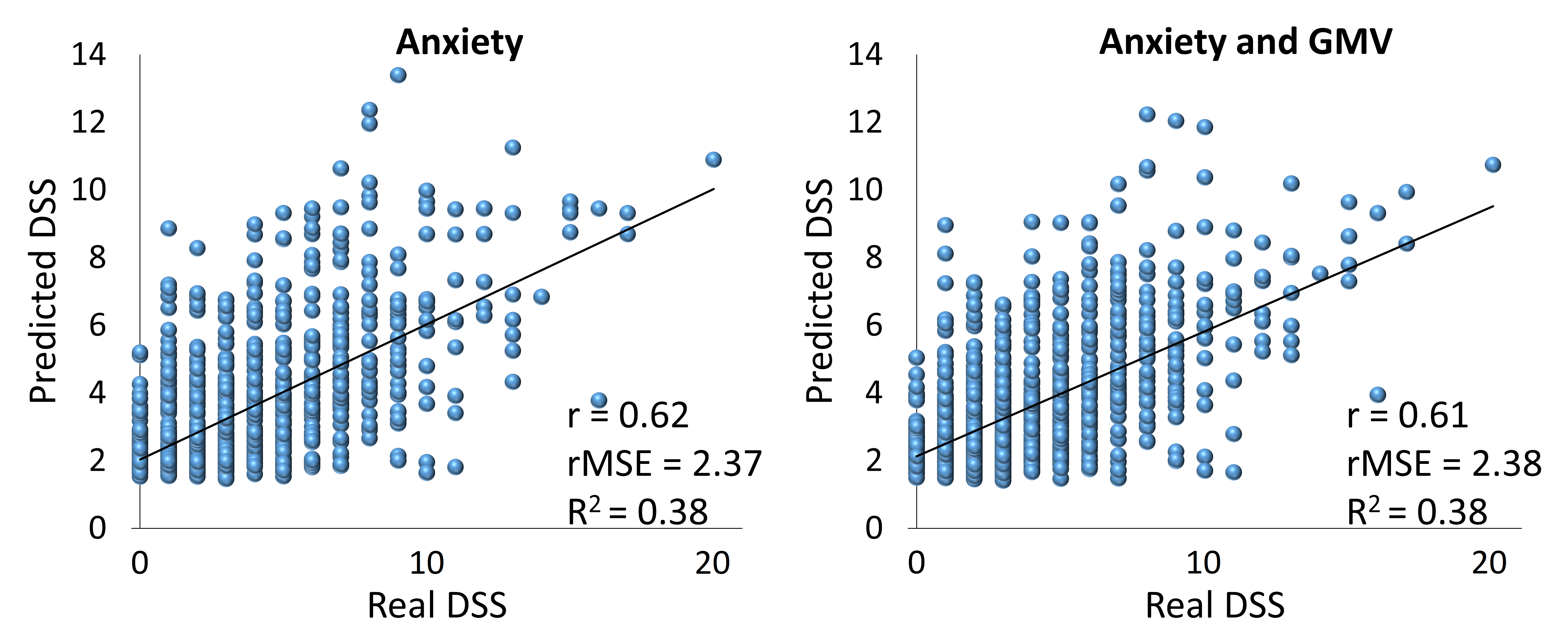
**eFigure 3.** Prediction of DSS in HCP-Young dataset based on only anxiety and a combination of anxiety and GMV. (DSS: depressive symptoms severity after excluding two sleep-related items, r: correlation coefficient between real and predicted DSS, GMV: gray matter volume, rMSE: root mean squared error, R^2^: determination coefficient).

Primarily based on the **eFigure 3**, it seems that the prediction of DSS based on anxiety scores is strong. While the correlation coefficient between DSS and anxiety scores itself is robust (r = 0.63), which means if we put features as the output (without any analyses) we would have better results than the ML results. Besides, a combination of anxiety and GMV features also could not predict DSS. Therefore, ML models were unable to predict DSS based on anxiety features.

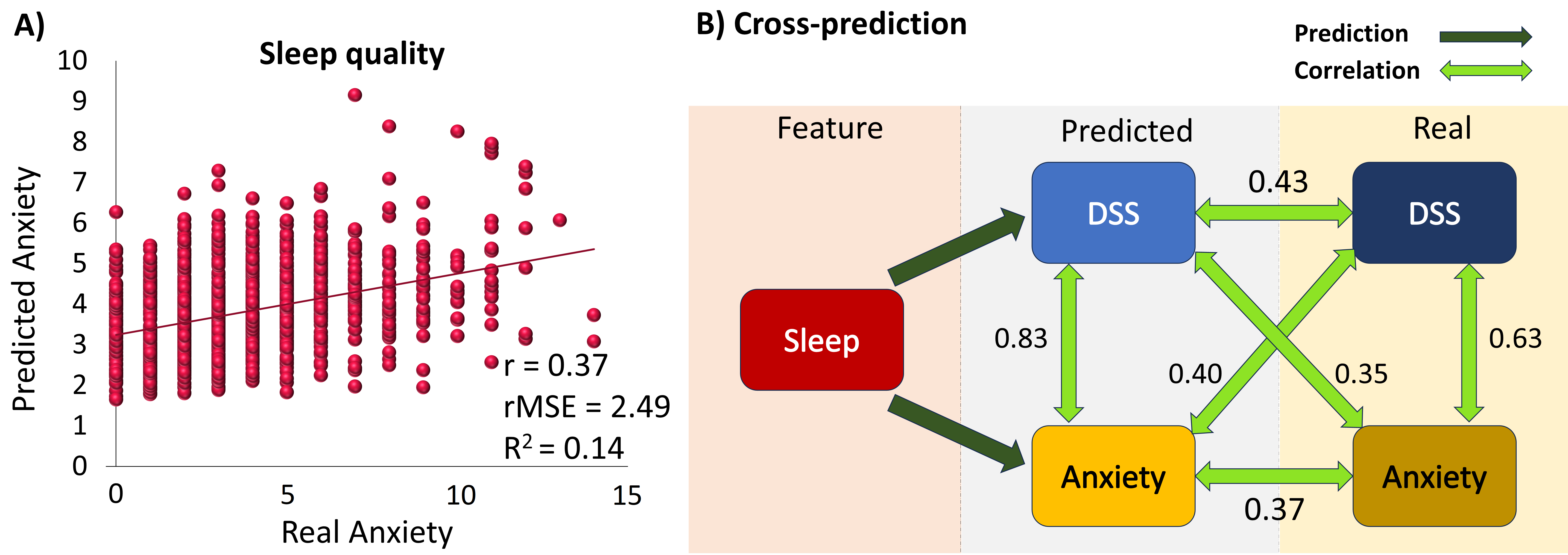

**eFigure 4.** Cross-prediction of anxiety and DSS in HCP-Young dataset. (DSS: depressive symptoms severity after excluding two sleep-related items, r: correlation coefficient between real and predicted anxiety, rMSE: root mean squared error, R^2^: determination coefficient).

To identify whether there is a collinearity between DSS and anxiety, we hypothesized that if the prediction model of DSS can predict DSS better than anxiety and also if the prediction model of anxiety can predict anxiety better than DSS, we can dedicate that there is no collinearity between DSS and anxiety. Hence, we predicted anxiety and DSS based on sleep quality and assessed if the correlation between predicted and real DSS was stronger than the correlation between predicted DSS and real anxiety and also if the correlation between real and predicted anxiety was stronger than the correlation between predicted anxiety and real DSS.

The results of cross-prediction show that the correlation between predicted and real DSS (r = 0.43) is more than the correlation between predicted DSS and real anxiety (r = 0.35) which confirms one of the two conditions of the hypothesis. Further, sleep quality predicted the amount of anxiety (r = 0.37) which was more correlated with DSS (r = 0.40). Therefore, we assessed the variance inflation factor (VIF) between variables. The VIF between sleep quality, DSS, and anxiety were 1.13, 1.68, and 1.72 respectively. All of the VIF scores are less than 3 and showed there is no collinearity between variables.

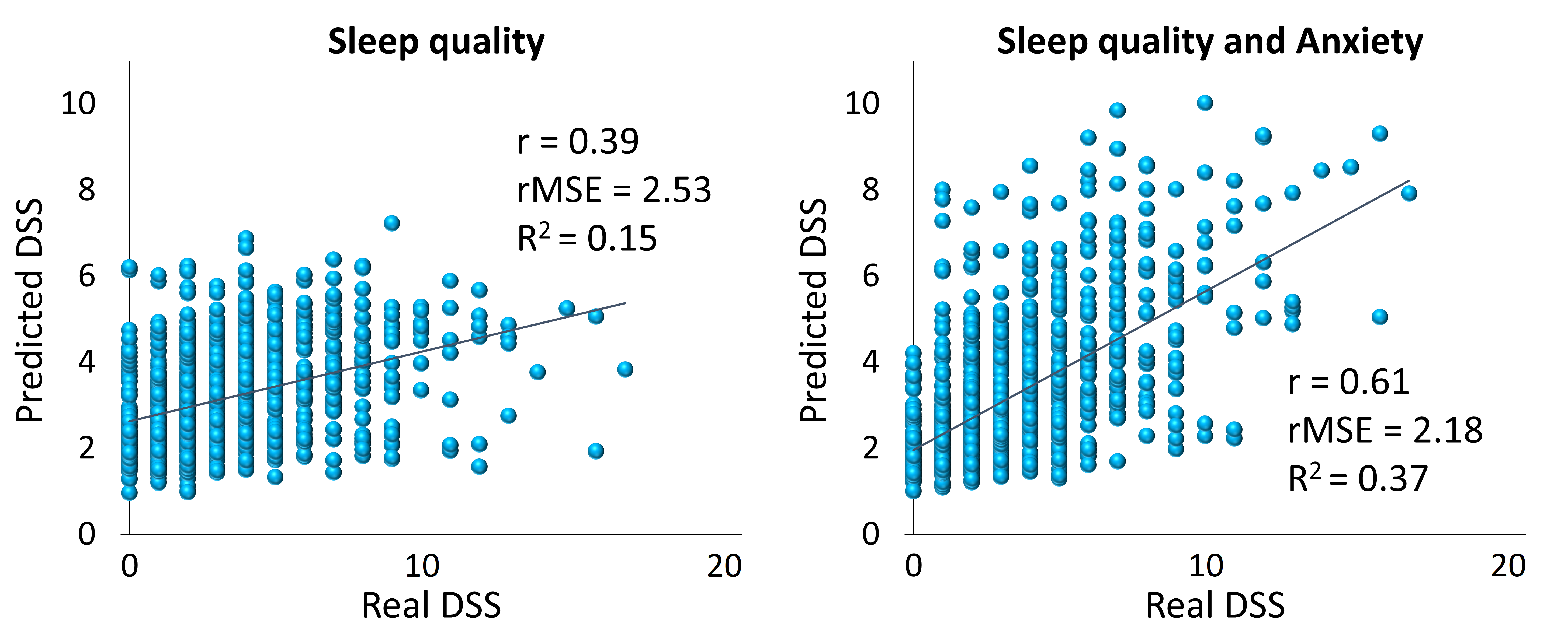

**eFigure 5.** Prediction of DSS in the HCP-Young dataset after excluding 103 participants who experienced at least one episode of depression. (DSS: depressive symptoms severity after excluding two sleep-related items, r: correlation coefficient between real and predicted DSS, rMSE: root mean squared error, R^2^: determination coefficient).

The results of prediction after excluding participants who had experienced at least one episode of depression remained robust and showed that ML models are responsible for general populations and healthy people.

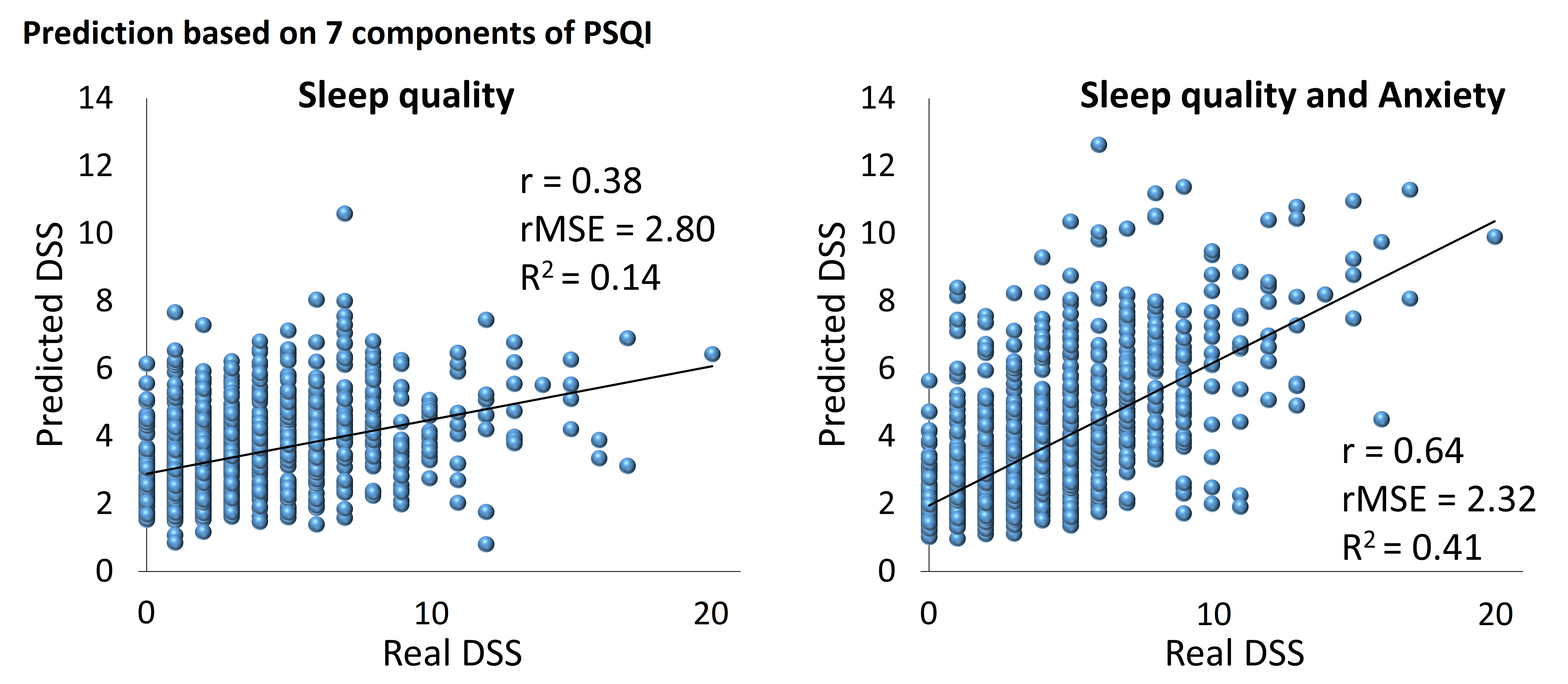
**eFigure 6.** Prediction of DSS based on seven components (subjective sleep quality, sleep latency, sleep duration, habitual sleep efficiency, sleep disturbances, use of sleep medicine, and daytime dysfunction) of PSQI in the HCP-Young dataset (PSQI: Pittsburgh Sleep Quality Index, DSS: depressive symptoms severity after excluding two sleep-related items, r: correlation coefficient between real and predicted DSS, rMSE: root mean squared error, R^2^: determination coefficient).

To identify the most important sleep quality parameter in the prediction of DSS we used seven components that identify the total sleep quality score that is calculated from the PSQI questionnaire to test prediction power at first (**eFigure 6**). Then, since the results were robust, we calculated the feature importance (**eFigure 7**) of this model.

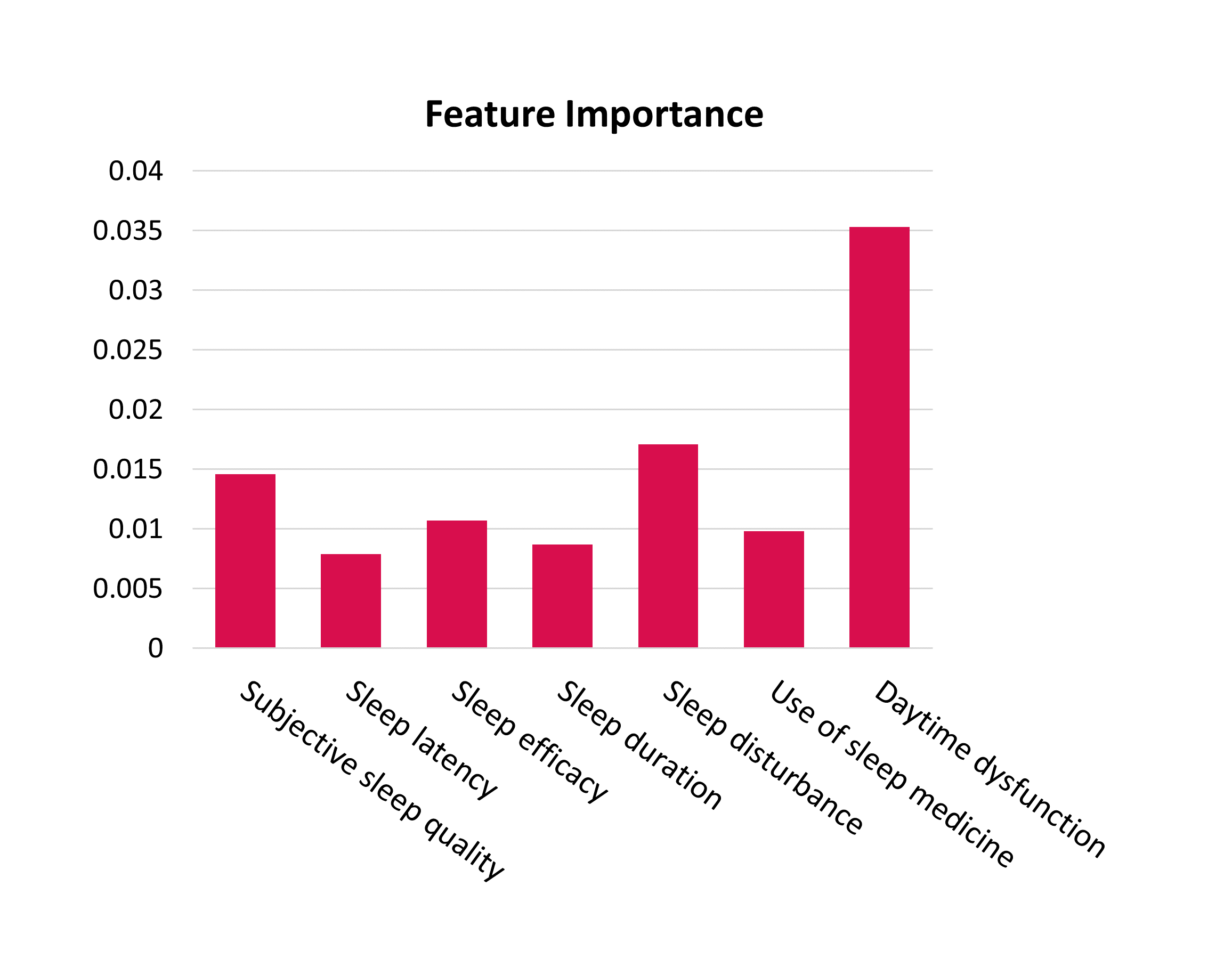

**eFigure 7.** Feature importance in prediction of DSS (depressive symptoms severity after excluding two sleep-related items) based on seven components of sleep quality.

The feature importance bar chart (**eFigure 7**) shows that the most important parameter which predicts DSS is dysfunction during the day that might come from sleep problems. At the second stage, different problems during the night that cause sleep disturbances (i.e., waking up at the middle of the night or feeling too cold/hot, etc.) are the most important predictors for DSS. The third important predictor of DSS is the participants’ perception of their sleep quality situation. Then, using sleep medicine, duration of sleep, and the time takes to fall asleep are the next priorities respectively.

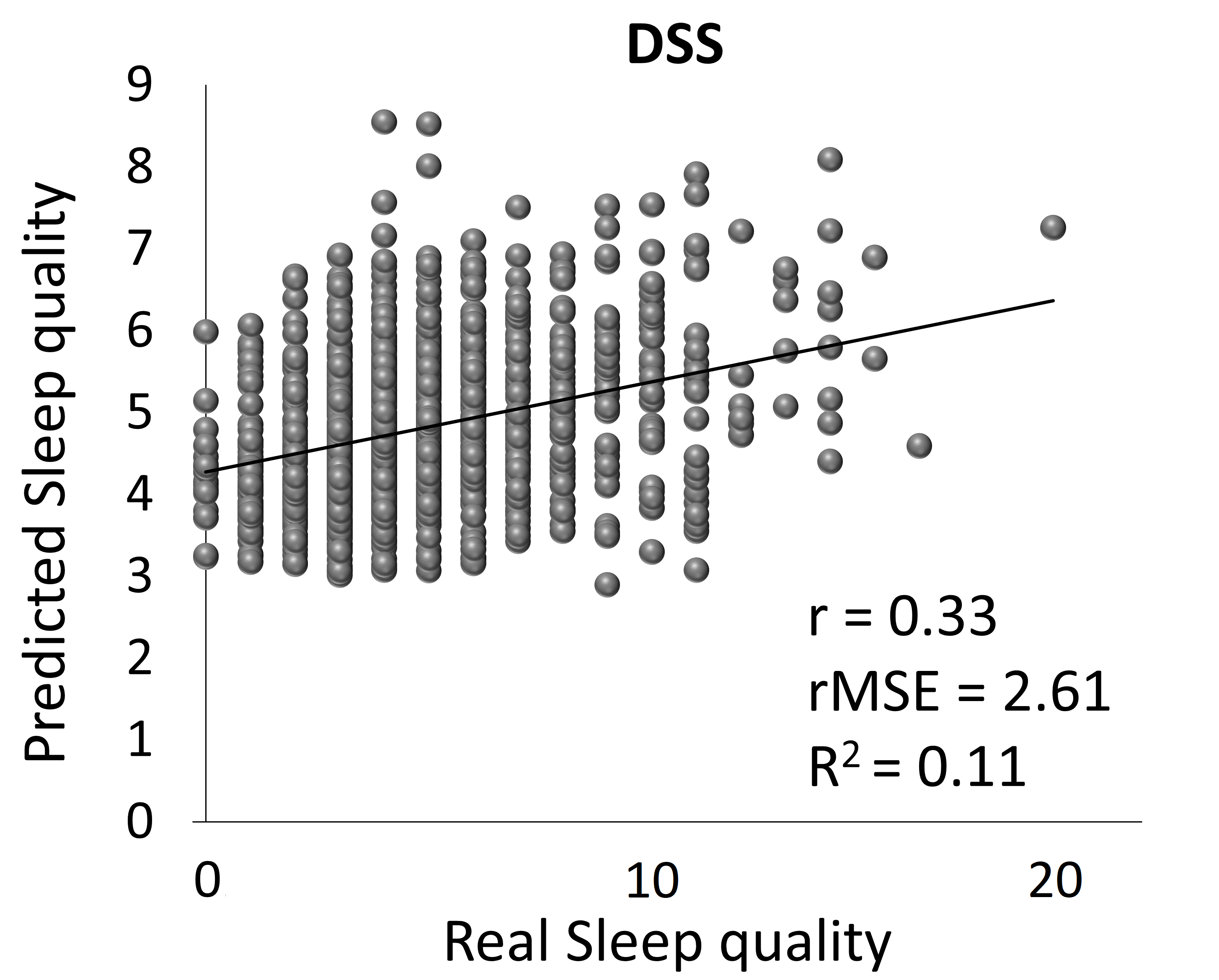

**eFigure 8.** Prediction of sleep quality based on DSS in HCP-Young dataset. (DSS: depressive symptoms severity after excluding two sleep-related items, r: correlation coefficient between real and predicted DSS, rMSE: root mean squared error, R^2^: determination coefficient).

Although DSS can predict sleep quality (r = 0.33, R^2^ = 0.11, rMSE = 2.61) (**eFigure 8**), its prediction power is significantly less than the reverse direction (r = 0.43, R^2^ = 0.18, rMSE = 2.73) (**Figure 2A**). prediction of sleep quality was performed using 12 depressive-related items of the Achenbach Adult Self-Report (ASR) for ages 18-59 (DSS items).

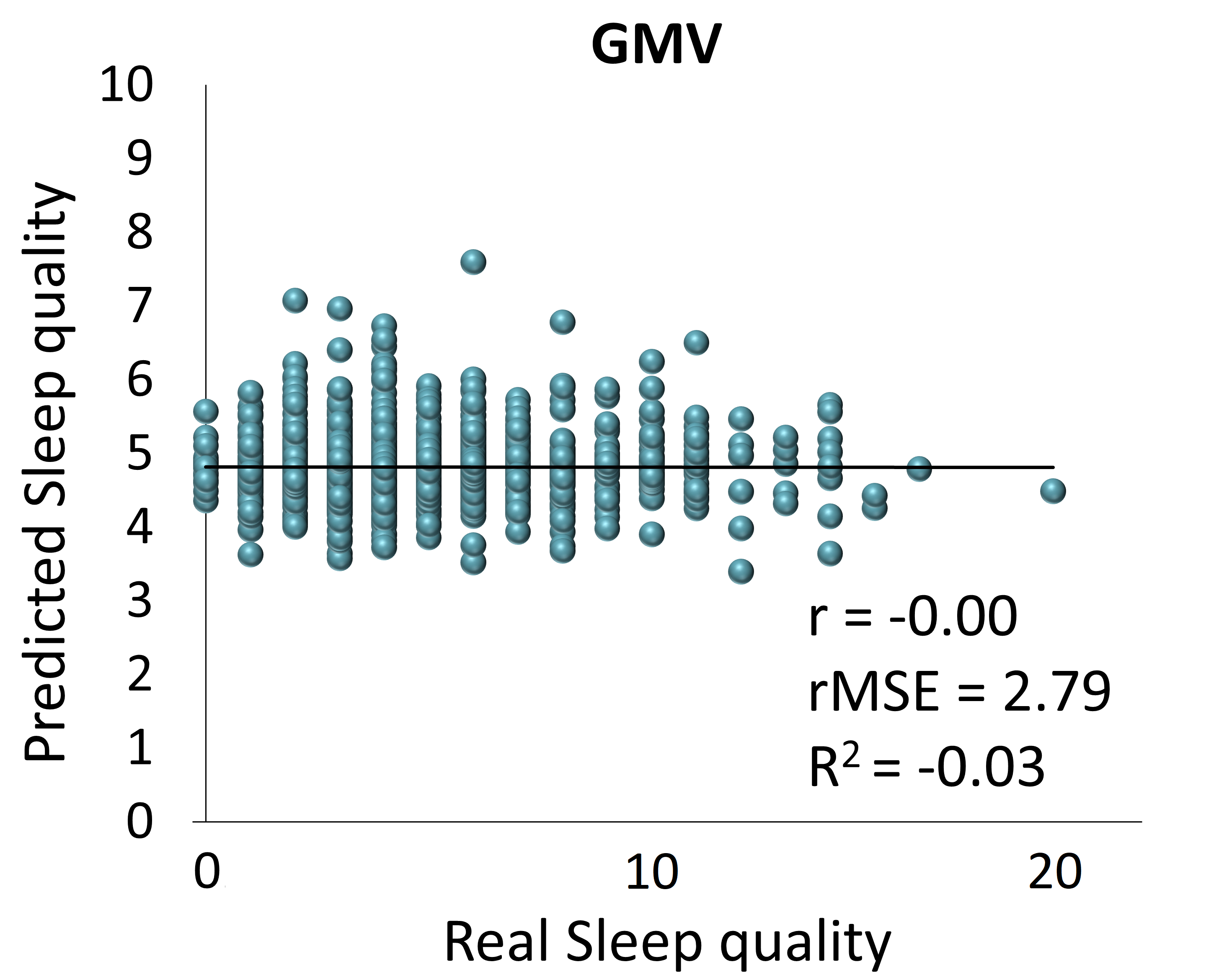

**eFigure 9.** Prediction of sleep quality based on GMV in the HCP-Young dataset. (GMV: gray matter volume, r: correlation coefficient between real and predicted sleep quality, rMSE: root mean squared error, R^2^: determination coefficient).

since there was a significant correlation between sleep quality and GMV (**eFigure 11**) we assessed whether GMV could predict sleep quality or not. The result of this analysis (**eFigure 9**) showed that GMV could not predict sleep quality either.

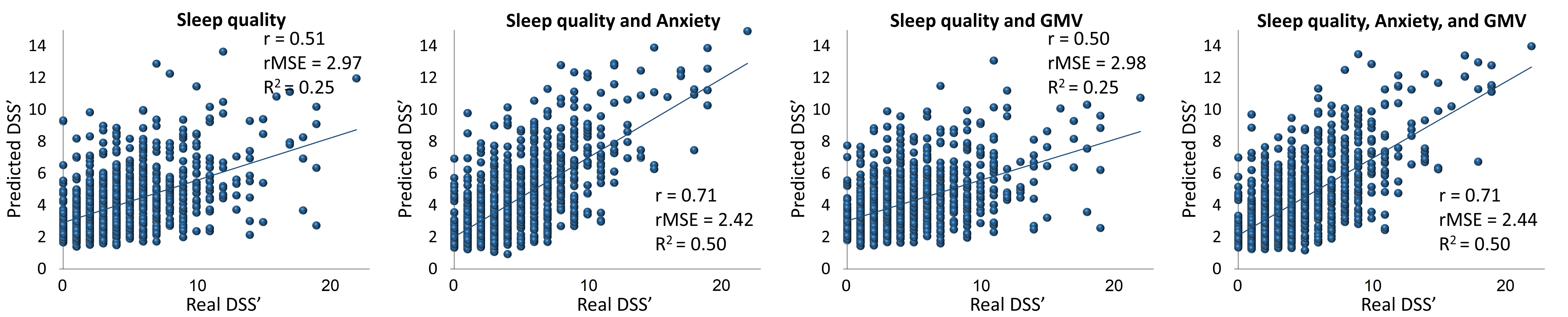

**eFigure 10.** Prediction of DSS’ in the HCP-Young dataset after including two sleep-related items of depressive symptoms severity (DSS’: depressive symptoms severity including two sleep-related items, GMV: gray matter volume, r: correlation coefficient between real and predicted DSS, rMSE: root mean squared error, R^2^: determination coefficient).

We aimed to assess when we include sleep-related scores of the DSS questionnaire (like other studies that have used depressive measures that had sleep-related items) and how much the prediction will improve. Hence, we included sleep-related items (8^th^ and 11^th^ questions of depressive related items) and made DSS’ and repeated the main analyses with this score. **eFigure 10** shows that this will improve the models’ performance. Therefore, we decided to compare DSS and DSS’ and assess how much of the relationship between DSS’ and sleep quality is because of these two sleep-related items of depressive problems (**eFigure 11**).

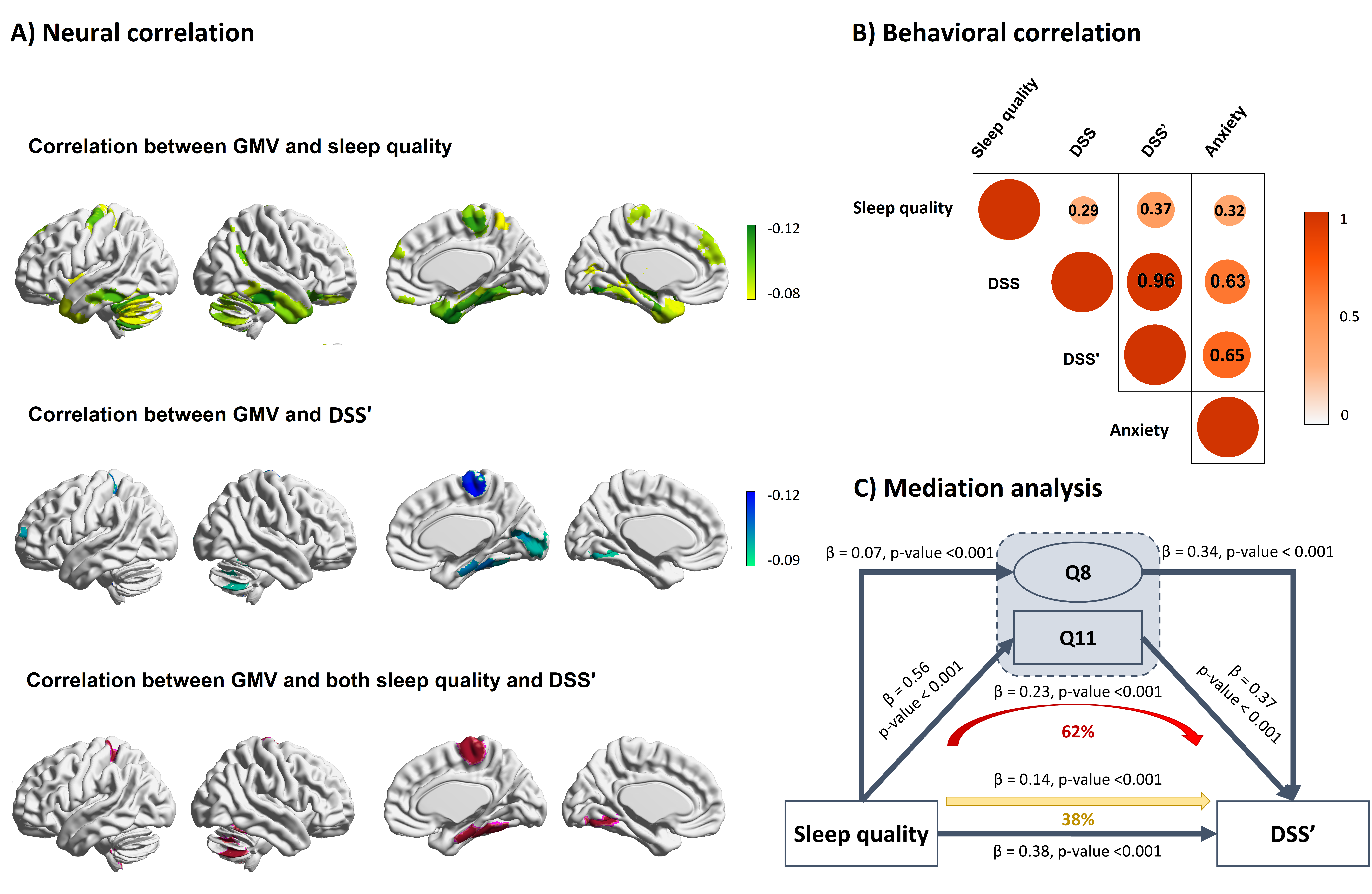

**eFigure 11.** Comparison of DSS and DSS’ in HCP-Young dataset. A) neural correlations we can find when we use DSS’. B) Correlations between phenotypic parameters used in this study. C) Mediational role of sleep related items of depressive problems in the relationship between sleep quality and DSS’. (DSS’: depressive symptoms severity including two sleep-related items, DSS: depressive symptoms severity after excluding two sleep-related items, GMV: gray matter volume, β: path coefficient, Q8 : 8th question of DSS’: “I sleep more than most other people during day and or night”, Q11 : 11th question of DSS’: “I have trouble sleeping”)

Although DSS and anxiety had no significant correlation with GMV, the results of analyses in **eFigure 11A**, show the brain areas (somatosensory area, primary motor cortex, extra-striate cortex, temporal lobe, parahippocampus, and cerebellum) in which their GMVs had significant correlation with both sleep quality and DSS’. So, these correlations between DSS’ and GMVs are because of two sleep-related items of DSS’ and generally is the correlation between these items and GMV. We assessed partial Pearson correlations between behavioral measures controlling for age, sex, and total GMV (**eFigure 11B**). Importantly, the result of mediation analysis (**eFigure 11C**) shows that 62% of covariance between sleep quality and DSS’ is because of sleep-related items of DSS’. Meanwhile, 90% of this effect is because of Q11 (11^th^ question of DSS’: “I have trouble sleeping”) and the other 10% is because of Q8 (8^th^ question of DSS’: “I sleep more than most other people during day and or night”).

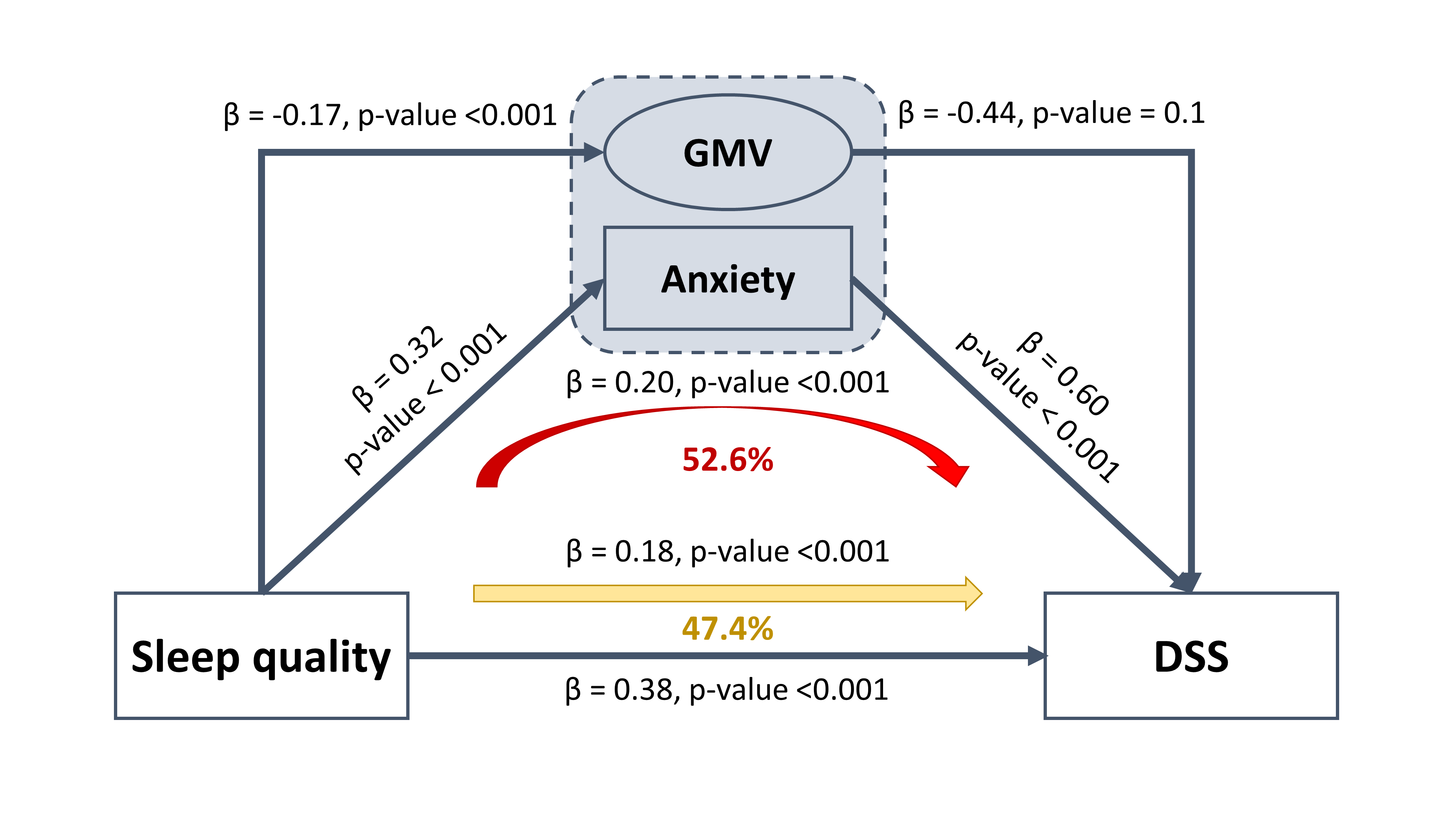

**eFigure 12.** Mediation analyses of GMV and anxiety. The standardized total effect of sleep quality on DSS is 0.38, and the direct effect is 0.18. The standardized indirect effect of sleep quality on DSS is 0.20. The direct effect of sleep quality on anxiety is 0.32, and on GMV is 0.-0.17. In addition, the direct effect of anxiety on DSS is 0.60, and the direct effect of GMV on DSS is -0.44, but it is insignificant. Hence, GMV has no mediatory role in the link between sleep quality and DSS, but anxiety has a partial mediatory role (52.6% of total effect size) in the link between sleep quality and DSS. (DSS: depressive symptoms severity, β: path coefficient).

After excluding sleep-related items of depressive problems scores (which had affected the amount of relationship (62% of covariance) between sleep quality and DSS’) we aimed to know the underlying mechanisms of the link between sleep quality and DSS. Comparing the results of mediational analyses (**eFigure 12**), we found anxiety problems score as a strong mediator of the link between sleep quality and DSS, while GMV has no mediatory role in the link between sleep quality and DSS (the p-value of the path coefficient of GMV on DSS is not significant p-value = 0.1).

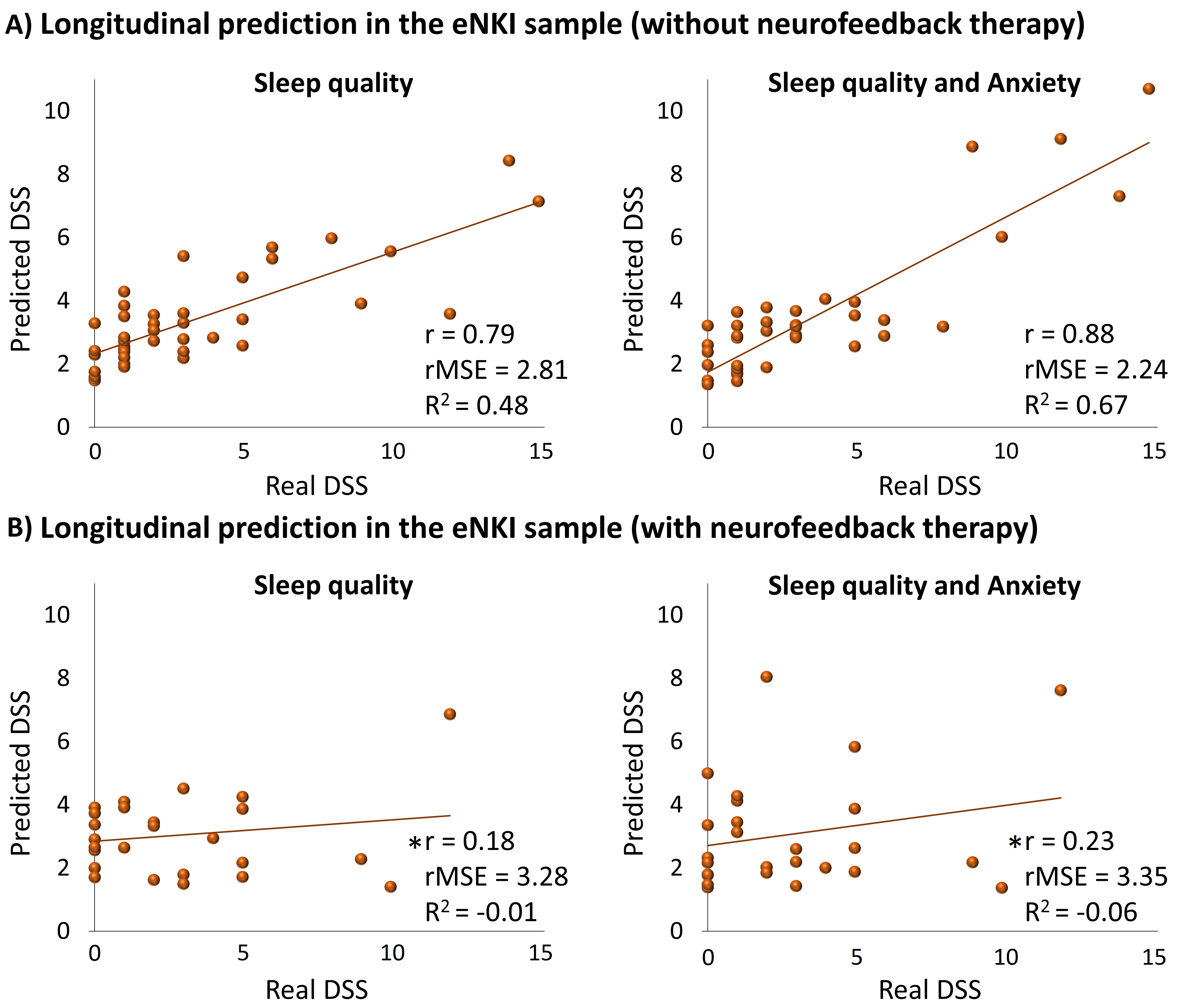

**eFigure 13.** Longitudinal prediction of DSS in the eNKI sample after separating subjects who had received neurofeedback therapy. A) prediction of DSS in the participants (N = 40) who had not received neurofeedback therapy between their first and second records (there were 1-4 years gap between the first and second records). B) prediction of DSS in the participants (N = 26) who had received neurofeedback therapy between their first and second records (there were 1-4 years gap between the first and second records). (DSS: depressive symptoms severity, r: correlation coefficient between real and predicted DSS, *: p-value > 0.1, rMSE: root mean squared error, R^2^: determination coefficient).

In the longitudinal analyses of predicting DSS, we found 66 participants who had longitudinal records, 40 of whom did not receive neurofeedback therapy, and the other 26 participants did not receive neurofeedback therapy. when we assessed participants who had not received neurofeedback therapy between their first and second records, prediction power significantly improved (**eFigure 13A**), and interestingly, the DSS of participants who had received neurofeedback therapy was not predictable (**eFigure 13B**). The average amount of DSS of participants who had received neurofeedback therapy was (mean _DSS_ = 2.85, std _DSS_ = 3.31) and the others were (mean _DSS_ = 3.40, std _DSS_ =3.90). These results collectively revealed the susceptibility of ML models to predict future DSS which can make the opportunity to prevent the severity of depressive symptoms by taking therapeutical interventions.

**References**

1.	Glasser MF, Sotiropoulos SN, Wilson JA, et al. The minimal preprocessing pipelines for the Human Connectome Project. *NeuroImage* 2013; **80**: 105-24.

2.	HCP 3T Imaging Protocol Overview. <https://protocols.humanconnectome.org/HCP/3T/imaging-protocols.html>.

3.	Salimi-Khorshidi G, Douaud G, Beckmann CF, Glasser MF, Griffanti L, Smith SM. Automatic denoising of functional MRI data: combining independent component analysis and hierarchical fusion of classifiers. *Neuroimage* 2014; **90**: 449-68.

4.	Mandal S RF, Sasse L, Komeyer V, Patil K, Hamdan S, Hoffstaedter F, Poldrack B, Weiss S. (2023). juaml/junifer: v0.0.3 (v0.0.3). Zenodo. <https://doi.org/10.5281/zenodo.8176570>.

5.	Cox RW. AFNI: software for analysis and visualization of functional magnetic resonance neuroimages. *Comput Biomed Res* 1996; **29**(3): 162-73.

6.	Zang Y, Jiang T, Lu Y, He Y, Tian L. Regional homogeneity approach to fMRI data analysis. *Neuroimage* 2004; **22**(1): 394-400.

7.	Zou QH, Zhu CZ, Yang Y, et al. An improved approach to detection of amplitude of low-frequency fluctuation (ALFF) for resting-state fMRI: fractional ALFF. *J Neurosci Methods* 2008; **172**(1): 137-41.

8.	Schaefer A, Kong R, Gordon EM, et al. Local-Global Parcellation of the Human Cerebral Cortex from Intrinsic Functional Connectivity MRI. *Cereb Cortex* 2018; **28**(9): 3095-114.

9.	Fan L, Li H, Zhuo J, et al. The Human Brainnetome Atlas: A New Brain Atlas Based on Connectional Architecture. *Cereb Cortex* 2016; **26**(8): 3508-26.

10.	Yeo BT, Krienen FM, Sepulcre J, et al. The organization of the human cerebral cortex estimated by intrinsic functional connectivity. *J Neurophysiol* 2011; **106**(3): 1125-65.

11.	Kira K, Rendell LA. The Feature Selection Problem: Traditional Methods and a New Algorithm. AAAI Conference on Artificial Intelligence; 1992; 1992.

12.	Arbuckle JL. Amos (Version 24.0) [Computer Program]. *Chicago: SPSS* 2019.

13.	Buysse DJ, Reynolds CF, Monk TH, Berman SR, Kupfer DJ. The Pittsburgh sleep quality index: A new instrument for psychiatric practice and research. *Psychiatry Research* 1989; **28**(2): 193-213.
